# Exome-wide association study to identify rare variants influencing COVID-19 outcomes: Results from the Host Genetics Initiative

**DOI:** 10.1101/2022.03.28.22273040

**Authors:** Guillaume Butler-Laporte, Gundula Povysil, Jack A. Kosmicki, Elizabeth T Cirulli, Theodore Drivas, Simone Furini, Chadi Saad, Axel Schmidt, Pawel Olszewski, Urszula Korotko, Mathieu Quinodoz, Elifnaz Çelik, Kousik Kundu, Klaudia Walter, Junghyung Jung, Amy D Stockwell, Laura G Sloofman, Daniel M. Jordan, Ryan C. Thompson, Diane Del Valle, Nicole Simons, Esther Cheng, Robert Sebra, Eric E. Schadt, Seunghee Schulze-Kim, Sacha Gnjatic, Miriam Merad, Joseph D. Buxbaum, Noam D. Beckmann, Alexander W. Charney, Bartlomiej Przychodzen, Timothy Chang, Tess D Pottinger, Ning Shang, Fabian Brand, Francesca Fava, Francesca Mari, Karolina Chwialkowska, Magdalena Niemira, Szymon Pula, J Kenneth Baillie, Alex Stuckey, Antonio Salas, Xabier Bello, Jacobo Pardo-Seco, Alberto Gómez-Carballa, Irene Rivero-Calle, Federico Martinón-Torres, Andrea Ganna, Konrad J Karczewski, Kumar Veerapen, Mathieu Bourgey, Guillaume Bourque, Robert JM Eveleigh, Vincenzo Forgetta, David Morrison, David Langlais, Mark Lathrop, Vincent Mooser, Tomoko Nakanishi, Robert Frithiof, Michael Hultström, Miklos Lipcsey, Yanara Marincevic-Zuniga, Jessica Nordlund, Kelly M. Schiabor Barrett, William Lee, Alexandre Bolze, Simon White, Stephen Riffle, Francisco Tanudjaja, Efren Sandoval, Iva Neveux, Shaun Dabe, Nicolas Casadei, Susanne Motameny, Manal Alaamery, Salam Massadeh, Nora Aljawini, Mansour S. Almutairi, Yaseen M. Arabi, Saleh A. Alqahtan, Fawz S. Al Harthi, Amal Almutairi, Fatima Alqubaishi, Sarah Alotaibi, Albandari Binowayn, Ebtehal A. Alsolm, Hadeel El Bardisy, Mohammad Fawzy, COVID-19 Host Genetics Initiative, DeCOI Host Genetics Group, GEN-COVID Multicenter Study (Italy), Mount Sinai Clinical Intelligence Center, GEN-COVID consortium (Spain), GenOMICC Consortium, Japan COVID-19 Task Force, Regeneron Genetics Center, Daniel H Geschwind, Stephanie Arteaga, Alexis Stephens, Manish J. Butte, Paul C. Boutros, Takafumi N. Yamaguchi, Shu Tao, Stefan Eng, Timothy Sanders, Paul J. Tung, Michael E. Broudy, Yu Pan, Alfredo Gonzalez, Nikhil Chavan, Ruth Johnson, Bogdan Pasaniuc, Brian Yaspan, Sandra Smieszek, Carlo Rivolta, Stephanie Bibert, Pierre-Yves Bochud, Maciej Dabrowski, Pawel Zawadzki, Mateusz Sypniewski, Elżbieta Kaja, Pajaree Chariyavilaskul, Voraphoj Nilaratanakul, Nattiya Hirankarn, Vorasuk Shotelersuk, Monnat Pongpanich, Chureerat Phokaew, Wanna Chetruengchai, Katsuhi Tokunaga, Masaya Sugiyama, Yosuke Kawai, Takanori Hasegawa, Tatsuhiko Naito, Ho Namkoong, Ryuya Edahiro, Akinori Kimura, Seishi Ogawa, Takanori Kanai, Koichi Fukunaga, Yukinori Okada, Seiya Imoto, Satoru Miyano, Serghei Mangul, Malak S Abedalthagafi, Hugo Zeberg, Joseph J Grzymski, Nicole L Washington, Stephan Ossowski, Kerstin U Ludwig, Eva C Schulte, Olaf Riess, Marcin Moniuszko, Miroslaw Kwasniewski, Hamdi Mbarek, Said I Ismail, Anurag Verma, David B Goldstein, Krzysztof Kiryluk, Alessandra Renieri, Manuel A.R. Ferreira, J Brent Richards

## Abstract

Host genetics is a key determinant of COVID-19 outcomes. Previously, the COVID-19 Host Genetics Initiative genome-wide association study used common variants to identify multiple loci associated with COVID-19 outcomes. However, variants with the largest impact on COVID-19 outcomes are expected to be rare in the population. Hence, studying rare variants may provide additional insights into disease susceptibility and pathogenesis, thereby informing therapeutics development. Here, we combined whole-exome and whole-genome sequencing from 21 cohorts across 12 countries and performed rare variant exome-wide burden analyses for COVID-19 outcomes. In an analysis of 5,085 severe disease cases and 571,737 controls, we observed that carrying a rare deleterious variant in the SARS-CoV-2 sensor toll-like receptor *TLR7* (on chromosome X) was associated with a 5.3-fold increase in severe disease (95% CI: 2.75-10.05, p=5.41×10^−7^). This association was consistent across sexes. These results further support *TLR7* as a genetic determinant of severe disease and suggest that larger studies on rare variants influencing COVID-19 outcomes could provide additional insights.

**Author Summary:** COVID-19 clinical outcomes vary immensely, but a patient’s genetic make-up is an important determinant of how they will fare against the virus. While many genetic variants commonly found in the populations were previously found to be contributing to more severe disease by the COVID-19 Host Genetics Initiative, it isn’t clear if more rare variants found in less individuals could also play a role. This is important because genetic variants with the largest impact on COVID-19 severity are expected to be rarely found in the population, and these rare variants require different technologies to be studies (usually whole-exome or whole-genome sequencing). Here, we combined sequencing results from 21 cohorts across 12 countries to perform a rare variant association study. In an analysis comprising 5,085 participants with severe COVID-19 and 571,737 controls, we found that the gene for toll-like receptor 7 (*TLR7*) on chromosome X was an important determinant of severe COVID-19. Importantly, despite being found on a sex chromosome, this observation was consistent across both sexes.

## Introduction

Despite successful vaccine programs, SARS-CoV-2 is still a major cause of mortality and widespread societal disruption^1,2^. While disease severity has correlated with well established epidemiological and clinical risk factors (e.g., advanced age, obesity, immunosuppression), these do not explain the wide range of COVID-19 presentations^3^. Hence, individuals without one of these known risk factors may have a genetic predisposition to severe COVID-19^4^. These genetic determinants to severe disease can, in turn, inform about the pathophysiology underlying COVID-19 severity and accelerate therapeutics development^5,6^.

Previous work on COVID-19 host genetics using genome-wide association studies (GWASs) revealed 23 statistically robust genetic loci associated with either COVID-19 severity or susceptibility^7–11^. Given that most GWASs use genetic data obtained from genome-wide genotyping followed by imputation to measure the association between a phenotype and genetic variation, their reliability and statistical power declines as a variant’s frequency decreases, especially at allele frequencies of less than 1%^12^. Ascertainment of rare genetic variation can be improved with sequencing technology^13^. Rare variants are expected to be enriched for larger effect sizes, due to evolutionary pressure on highly deleterious variants, and may therefore provide unique insights into genetic predisposition to COVID-19 severity. Identifying such genes may highlight critical control points in the host response to SARS-CoV-2 infection.

Measuring the effect of rare genetic variants on a given phenotype (here COVID-19) is difficult. Specifically, while variants of large effect on COVID-19 are more likely to be rare, the converse is not true, and most rare variants are not expected to strongly impact COVID-19 severity^14^. Therefore, unless large sample sizes and careful statistical adjustments are used, most rare variant genetic associations studies risk being underpowered, and are at higher risk of false or inflated effect estimates if significant associations are found between COVID-19 and genetic loci. This is exemplified by the fact that several rare variant associations reported for COVID-19 have not been replicated in independent cohorts^15–17^.

Here, we investigated the association of rare genetic variants on the risk of COVID-19 by combining gene burden test results from whole exome and whole genome sequencing. We build off recent work on exome-wide analyses^17^ and include close to 5 times the number of severe cases, with a more genetically diverse cohort, to better study the effect of rare variants on COVID-19. To our knowledge, this is the first rare genetic variant burden test meta-analysis ever performed on a worldwide scale, including 21 cohorts, in 12 countries, including all main continental genetic ancestries.

## Results

### Study population and outcome

The final analysis included up to 28,159 individuals infected with SARS-CoV-2, and up to 597,165 controls from 21 cohorts in 12 countries (**Figure 1**). Most participants were of European genetic ancestry (n=576,389), but the consortium also included participants of Admixed American (n=4,529), African (n=25,465), East Asian (n=4,716), Middle Eastern (n=4,977) and South Asian ancestries (n=9,943). These resulted in a genetically diverse sample of participants (**Figure 2**). Participating cohorts enrolled patients based on local protocols, and both retrospective and prospective designs were used. Genetic sequencing was also performed locally, and cohorts were provided with a specific framework for quality control analyses, but each were allowed to deviate based on individual needs. Both exome (n = 11 cohorts) and genome sequencing (n = 10 cohorts) were included in the meta-analyses. The mean age of participants was 55.6 years, and 55.9% were females.

**Figure 1:**
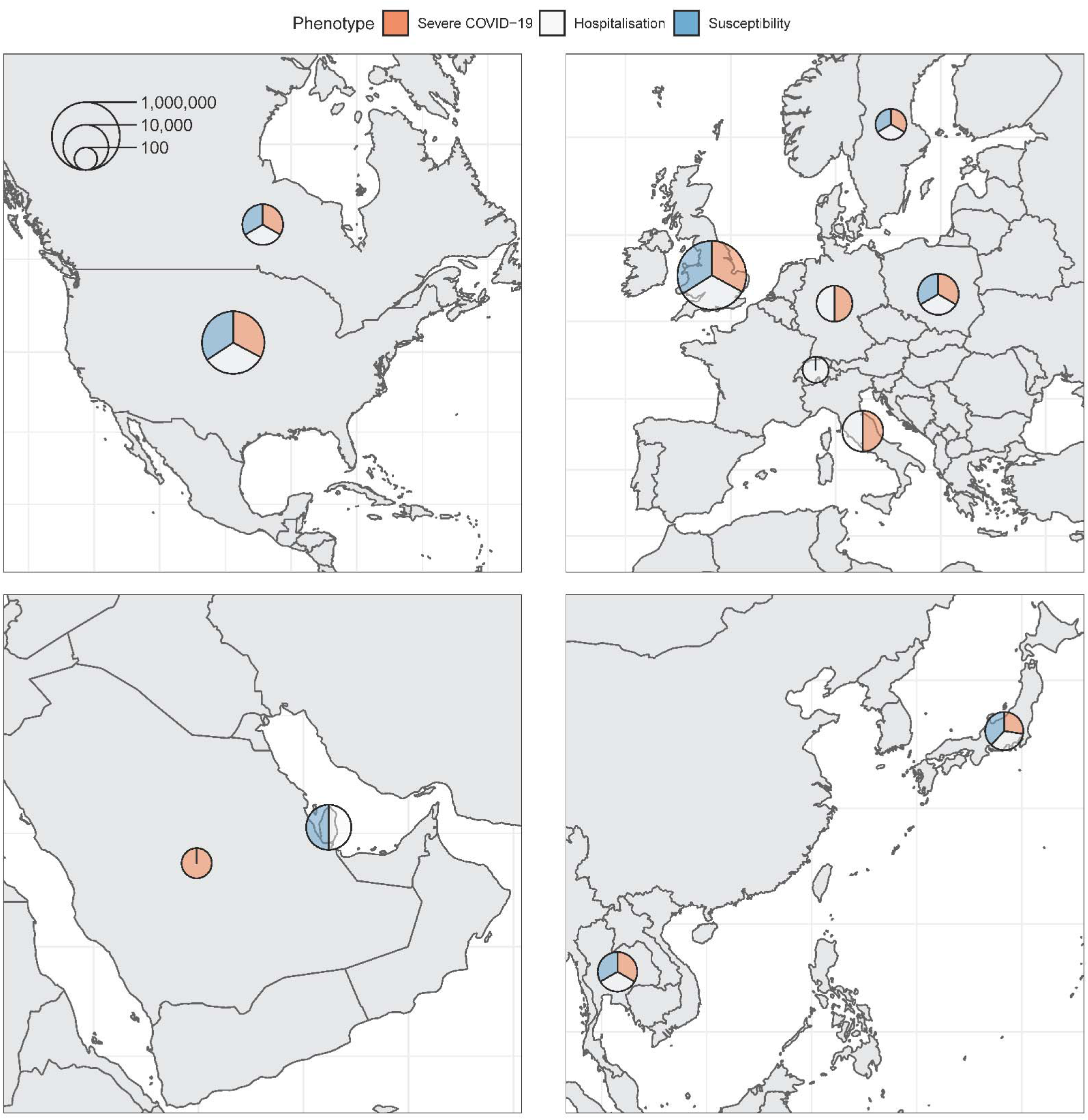
Maps of countries contributing data to the consortium. Sample sizes (cases and controls) for each phenotype were added and represented on the logarithmic scale by each circle. Relative contribution to each phenotype is represented by the three colors.

**Figure 2:**
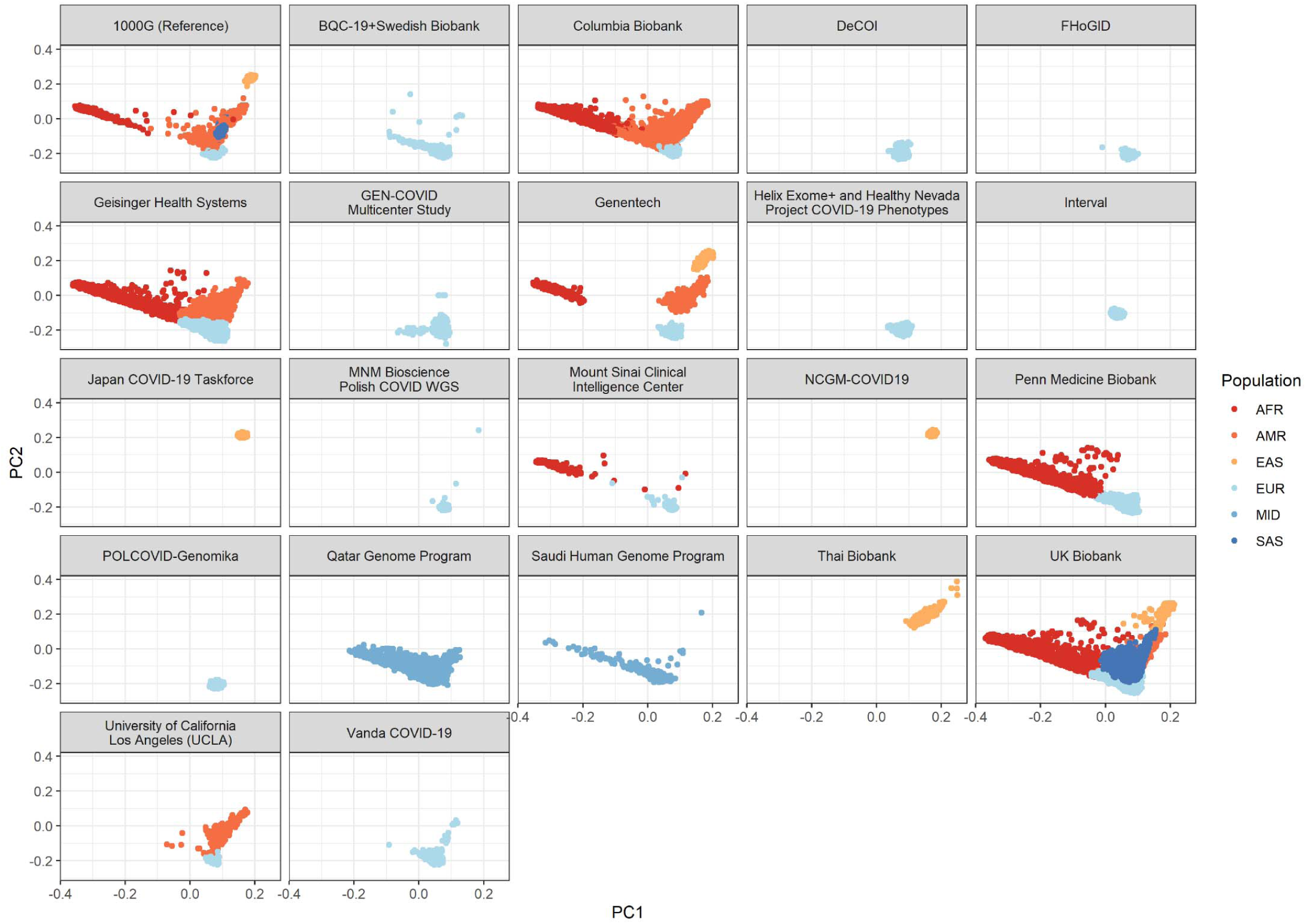
Participant’s genome projection on the first and second genetic principal components of the 1000G reference panel. AFR: African ancestry. AMR: admixed American ancestry. EAS: east Asian ancestry. EUR: European ancestry. MID: middle eastern ancestry. SAS: south Asian ancestry.

We studied three separate outcome phenotypes, as previously described by the COVID-19 Host Genetics Initiative (COVID-19 HGI)^8^. Briefly, the outcome cases were defined according to three standard COVID-19 HGI outcomes: A) severe disease: individuals with SARS-CoV-2 infection who died or required invasive respiratory support (extracorporeal membrane oxygenation, intubation with mechanical ventilation, high-flow oxygen support, or new bilevel or continuous positive airway pressure ventilation), B) hospitalisation: individuals with SARS-CoV-2 who died or required hospitalisation, and C) susceptibility to infection: any individual with SARS-CoV-2 infection. These are also referred to as A2, B2, and C2, respectively, in the COVID-19 HGI meta-analyses^8^. For all three phenotypes, controls were all individuals not classified as cases (including population controls with unknown COVID-19 status). The final meta-analyses included up to 5,085 cases and 571,737 controls for the severe disease outcome, 12,304 cases and 590,151 controls for the hospitalisation outcome, and 28,196 cases and 597,165 controls for the susceptibility outcome.

### Single-variant analysis

We first performed an exome-wide association study using single variants with a MAF (minor allele frequency) higher than 0.1% and an allele count of 6 or more in at least one cohort, with the same additive model and covariates used in the COVID-19 HGI GWAS^8^. Analyses were performed separately by each cohort and each ancestry using Firth regression as applied in the Regenie software^18^. Firth regression is a penalized likelihood regression method that provides unbiased effect estimates even in highly unbalanced case-control analyses^19^. The summary statistics were then meta-analyzed with a fixed effect inverse-variance weighted model within each ancestry, and then with a DerSimonian-Laird random effect model across ancestries.

The previously described Neanderthal chromosome 3 locus associated with COVID-19 outcomes^8,20^ was also found in all three phenotypes (**Figure 3, Supp. Figure 1**), with lead variants in the *CXCR6* gene for the severe COVID-19 phenotype (rs13059238), and in *FYCO1* in the hospitalisation phenotype (rs13069079), and for the *LIMD1* gene in the susceptibility phenotype (rs141045534). Reassuringly, each cohort provided summary statistics in the chromosome 3 locus, suggesting that the QC process was working as intended (allowing for sample sizes and number of cases) (**Supp Figures 2-4**).

**Figure 3:**
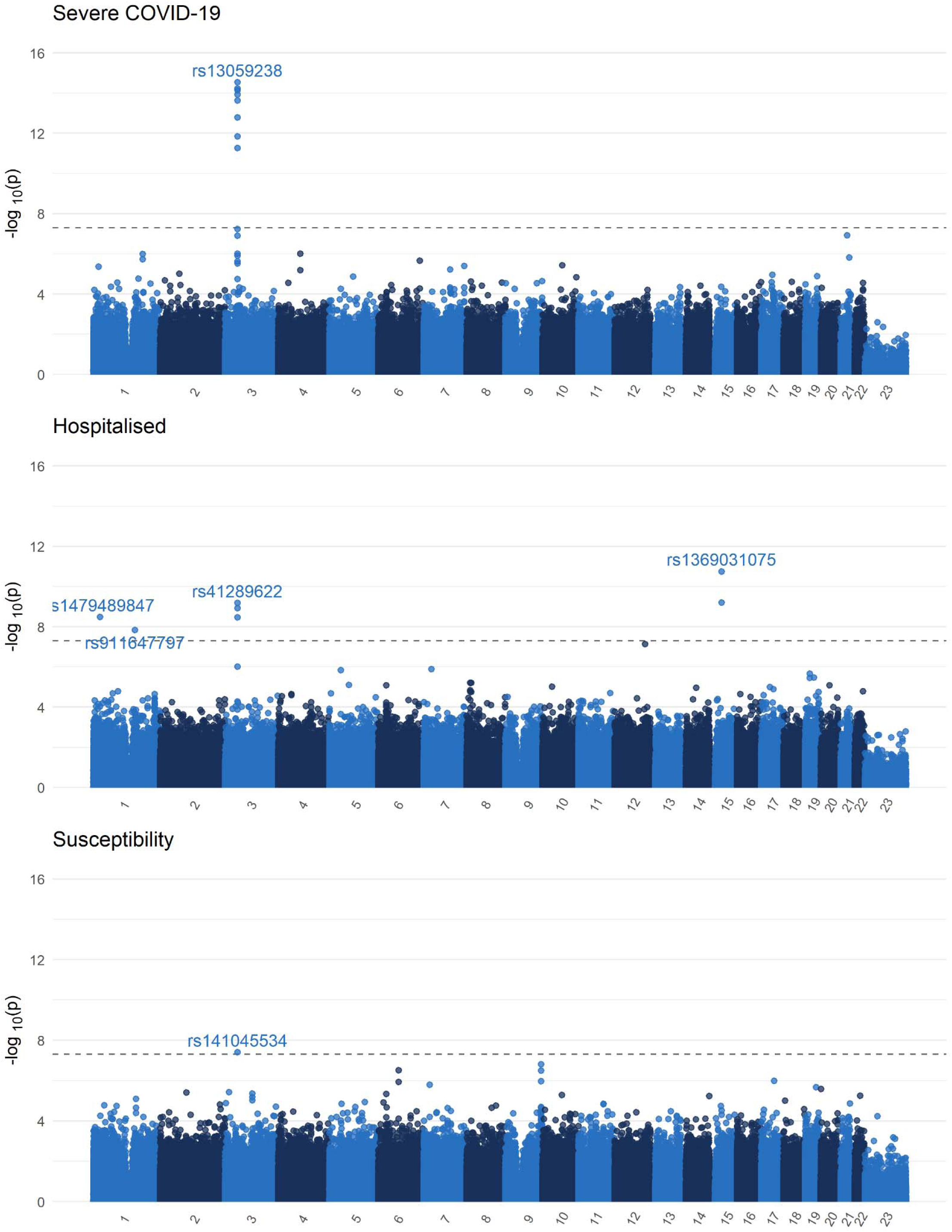
Single variant exome-wide association study Manhattan plot (MAF>0.1%). QQ-plot available in the **Supp. Figures**. Black dashed line demarcates the genome-wide significance threshold (p < 5×10^−8^).

Three other loci were found for the hospitalization phenotypes. One at *SRRM1* (rs1479489847, OR: 4.17, 95% CI 2.60-6.70, p=3.25×10^−9^), *IL6R* (rs911647797, OR: 6.19, 95% CI: 3.29-11.6, p=1.45×10^−8^), and another at cytoskeleton *FRMD5* (rs1369031075, OR: 4.06, 95% CI: 2.70-6.11, p=1.75×10^−11^). While these loci may hold biological plausibility (especially *IL6R*, given the use of IL-6 receptor inhibitors in the treatment of COVID-19), these associations were driven by two smaller cohorts (Genentech and Vanda, **Supp. Figure 5**). However, the *SRMM1* locus is located between two stretches of T nucleotides, while both the *FRMD5* and the *IL-*6 loci are within GC rich regions, making variant calling difficult. Hence, these findings will require validation, despite the biological plausibility.

Finally, all genetic inflation factors were below 1 (**Supp. Table 8**). Summary statistics for genome-wide significant variants can be found in **Supp. Table 9** and QQ-plots can be found in **Supp. Figure 1**.

### Burden test definition

Given the expected paucity of large-effect size rare deleterious variants, strategies have been devised to increase statistical power to test associations between rare variants and biomedically-relevant outcomes. One such strategy is to use burden tests^21^, where each variant is collapsed into larger sets of variants, and association is tested between groups of variants and an outcome. Here, we collapsed deleterious variants in each gene and devised the following burden test: for each gene, an individual received a score of 0 if they do not carry any deleterious variant, a score of 1 if they carry at least one non-homozygous deleterious variant, and a score of 2 if they carry at least 1 homozygous deleterious variant. As defined in previous studies on burden testing of rare variants^17,22^ deleterious variants were chosen using three masks: 1) “M1” which uses only predicted loss of function variants, 2) “M3” which uses all variants in M1, as well as indels of moderate consequence as predicted by Ensembl^23^, and missense variants classified as deleterious in 5 *in-silico* algorithms (see **Methods**), and 3) “M4”, which uses all variants in M1 and M3, and also adds all missense variants classified as deleterious in at least 1 of the *in-silico* algorithms.

The analyses were performed separately both for variants with MAF of less than 1%, and for variants of MAF less than 0.1%. We defined MAFs based on a combination of gnomAD^24^ MAF annotations, and of cohort-specific common variant exclusion lists. These common variant exclusion lists included variants that achieved a MAF of >1 % or >0.1 % in at least one study population within the consortium. To reduce the effect of fluctuations due to sampling, a minor allele count (MAC) ≥ 6 in the corresponding study was required for inclusion in the common variant list. Such “blacklists” have been shown to increase statistical power by removing variants at lower risk of being highly deleterious, and it reduces the risk of having cohort-specific false-positive variants being retained on the overall analysis^25^. Hence for each MAF threshold, each cohort removed any variant with a MAF above the threshold in either gnomAD or the corresponding common variant exclusion list.

The resulting score (either 0, 1, or 2) for each mask was then regressed on each of our three phenotypes using logistic regression, controlling for age, age^2, sex, sex*age, sex*age^2^, and 10 common variant (MAF > 1%) genetic principal components (the same covariates as for COVID-19 HGI GWASs^7,8^). Additionally, given that population genetic structure and its confounding effect on phenotypes is different at the rare variant level^26^, we also used the first 20 genetic principal components from rare variants (MAF<1%) as covariates in all our analyses. Analyses were otherwise done using the same approach as for single-variant analyses.

### Exome-wide burden test analyses results

Our meta-analysis included a total of 18,883 protein-coding genes, and all burden test genetic inflation factors, for all masks, were less than 1 (**Supp. Table 8**), suggesting that our results were not biased by population stratification and that Firth regression adequately adjusted for unbalanced case-control counts. Using an exome-wide significance p-value threshold of 0.05/20,000 = 2.5×10^−6^, we found 3 genes associated with one of the COVID-19 phenotypes in at least one mask in our meta-analyses (**Table 1, Supp. Figures 6-12**). Of specific interest, we observed that carrying a predicted loss of function or *in-silico* highly deleterious missense variant (i.e., mask M3) in the toll-like receptor 7 (*TLR7*) gene was associated with a 5.3-fold increase (95% CI: 2.7-10.1, p=5.41×10^−7^) in odds of severe COVID-19. *TLR7* is an important part of the innate viral immunity, encoding a protein that recognizes coronaviruses and other single-stranded RNA viruses, leading to upregulation of the type-1 and type-2 interferon pathway^27^. Results from the severe COVID-19 outcome analyses of *TLR7* with other masks also nearly reach our statistical significance threshold, with larger effects found in the M1 mask (OR: 13.6, 95% CI: 4.41-44.3, p=1.64×10^−5^) and smaller effect in the M4 mask (OR: 3.12, 95% CI: 1.91-5.10, p=5.30×10^−6^), though the latter was balanced by smaller standard errors due to the larger number of cases (3275 cases in M4 vs 1577 in M1), as expected. These findings further support previous reports of *TLR7* errors of immunity underlying severe COVID-19 presentations^17,28–31^.

**Table 1:** Exome-wide significant findings, as well as other *TLR7* results (for the severe phenotype only). Note that for Masks M1, all deleterious variants had a MAF<0.1%, and hence both burden tests (MAF<1% and 0.1%) gave the same results. Full results available in **Supp. Table 9**.

| Gene | Mask | Phenotype | MAF | Beta | Standard Error | Odds Ratio | 95% Confidence Interval | P-value | Heterogeneity p-value | N Cases 0\|1\|2 Burden Test | N Controls 0\|1\|2 Burden Test |
| --- | --- | --- | --- | --- | --- | --- | --- | --- | --- | --- | --- |
| Meta-Analysis Across Ancestries |  |  |  |  |  |  |  |  |  |  |  |
| MARK1 | M1 | Severe COVID-19 | <0.1% | 3.17 | 0.67 | 23.9 | 6.5-88.2 | 1.89x10<sup>-6</sup> | 0.883 | 1935\|4\|0 | 540031\|92\|0 |
| MARK1 | M1 | Severe COVID-19 | <1% | 3.17 | 0.67 | 23.9 | 6.5-88.2 | 1.89x10<sup>-6</sup> | 0.883 | 1935\|4\|0 | 540031\|92\|0 |
| MARK1 | M1 | Hospitalisation | <0.1% | 2.51 | 0.48 | 12.3 | 4.8-31.2 | 1.43x10<sup>-7</sup> | 0.893 | 6132\|8\|0 | 547943\|93\|0 |
| MARK1 | M1 | Hospitalisation | <1% | 2.51 | 0.48 | 12.3 | 4.8-31.2 | 1.43x10<sup>-7</sup> | 0.893 | 6132\|8\|0 | 547943\|93\|0 |
| RILPL1 | M1 | Severe COVID-19 | <0.1% | 3.01 | 0.64 | 20.2 | 5.8-70.7 | 2.42x10<sup>-6</sup> | 0.941 | 1745\|4\|0 | 558448\|121\|0 |
| TLR7 | M3 | Severe COVID-19 | <0.1% | 1.66 | 0.33 | 5.25 | 2.75-10.05 | 5.41x10<sup>-7</sup> | 0.755 | 3101\|2\|5 | 519047\|83\|47 |
| TLR7 | M3 | Severe COVID-19 | <1% | 1.63 | 0.33 | 5.10 | 2.67-9.72 | 7.48x10<sup>-7</sup> | 0.760 | 3275\|2\|5 | 519834\|85\|47 |
| Other TLR7 results for severe phenotype |  |  |  |  |  |  |  |  |  |  |  |
| TLR7 | M1 | Severe COVID-19 | <0.1% | 2.61 | 0.60 | 13.6 | 4.14-44.4 | 1.64x10<sup>-5</sup> | 0.820 | 1577\|0\|2 | 508987\|13\|11 |
| TLR7 | M1 | Severe COVID-19 | <1% | 2.61 | 0.60 | 13.6 | 4.14-44.4 | 1.64x10<sup>-5</sup> | 0.820 | 1577\|0\|2 | 508987\|13\|11 |
| TLR7 | M4 | Severe COVID-19 | <0.1% | 1.14 | 0.25 | 3.12 | 1.91-5.10 | 5.30x10<sup>-6</sup> | 0.854 | 3275\|3\|7 | 519616\|210\|139 |
| TLR7 | M4 | Severe COVID-19 | <1% | 1.11 | 0.24 | 3.03 | 1.90-4.85 | 3.43x10<sup>-6</sup> | 0.956 | 3273\|5\|8 | 521166\|221\|144 |

In the meta-analyses, we also found that pLoFs (M1) in *MARK1* were associated with a 23.9-fold increase in the odds of severe COVID-19 (95% CI: 6.5-88.2, p=1.89×10^−6^), and a 12.3-fold increase in the odds of hospitalisation due to COVID-19 (95% CI: 4.8-31.2, p=1.43×10^−7^). While the number of *MARK1* pLoFs (M1) found in severe and hospitalized cases was small (MAC=4 and MAC=8, respectively), the signal was consistent in our three largest cohorts: UK Biobank, Penn Medicine, and Geisinger Health Services (**Supp. Figures 9-12**). *MARK1* is a member of the microtubule affinity-regulating kinase family, and is involved in multiple biological processes, chief among which is the promotion of microtubule dynamics^32^. *MARK1* has previously been shown to interact with the SARS-CoV-2 ORF9b protein^33^, further supporting its potential role in COVID-19. Lastly, our meta-analyses also found marginal evidence for an association between severe COVID-19 and pLoFs (M1) in *RILPL1* (OR: 20.2, 95% CI: 5.8-70.7, p=2.42×10^−6^), a gene that, like *MARK1*, is associated with microtubule formation and ciliopathy^34^.

We then meta-analyzed p-values using the aggregated Cauchy association test^35^ (ACAT). ACAT accounts for correlation between test statistics (as is expected here) by treating p-values as Cauchy random variables, and taking their weighted average, which also is Cauchy distributed. With ACAT, the association between *TLR7* and severe COVID-19 (p=1.58×10^−6^), and between *MARK1* and hospitalisation (p=4.30×10^−7^) remained exome-significant (**Figure 4**). Full summary statistics are available in **Supp. Table 10**.

**Figure 4:**
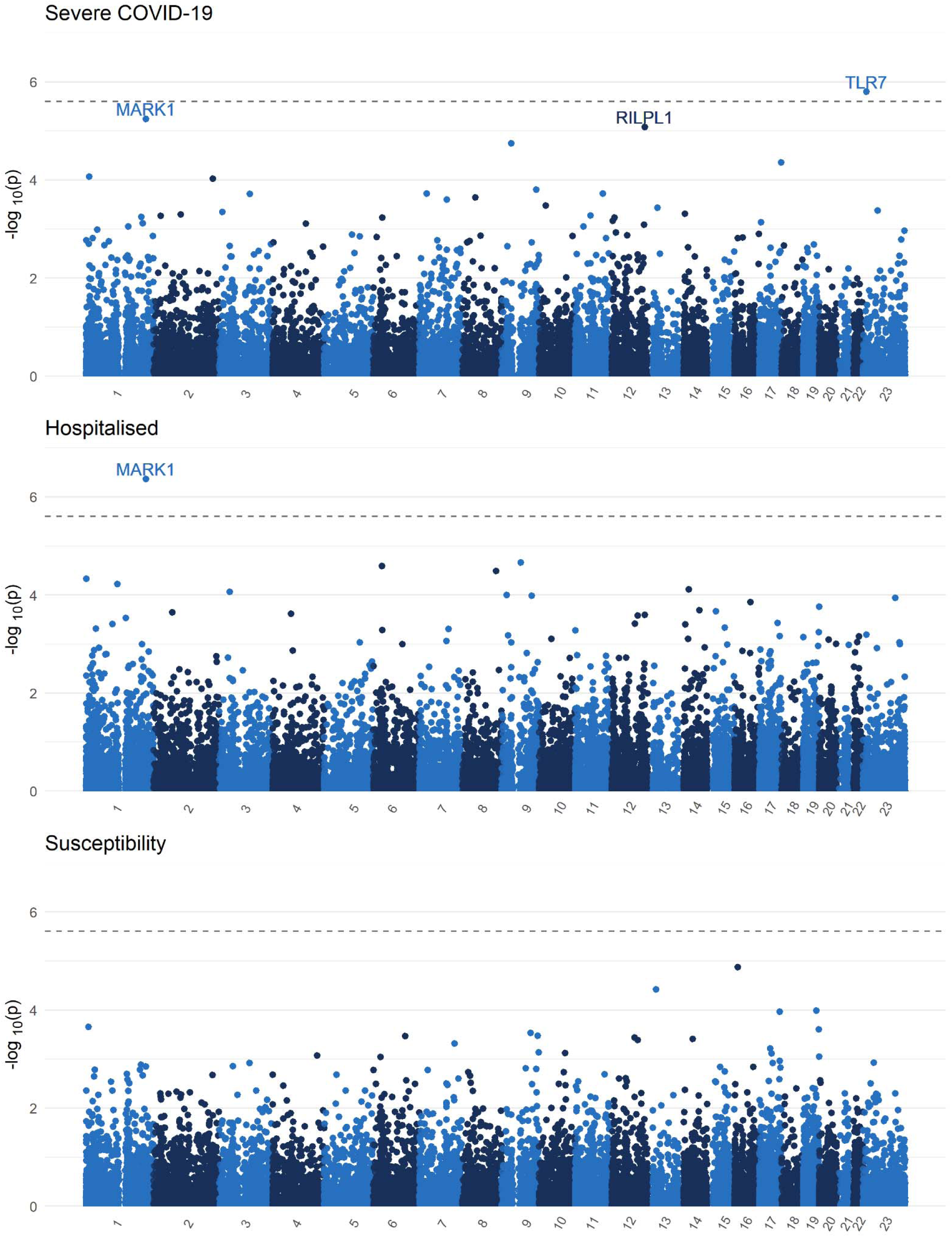
Exome burden test ACAT p-value meta-analysis Manhattan plots and QQ plots. QQ-plot available in the **Supp. Figures**. Black dashed line demarcates the Bonferroni significance threshold (p < 0.5/20,000).

Finally, we note that for both *TLR7* and *MARK1*, the signal was driven by European ancestry participants. Further, while the larger biobanks contributed to these findings, smaller prospective cohorts also provided cases with rare variants at both genes, highlighting the importance of study design in rare variant association testing (**Supp. Figures 6-12**).

### TLR7 sex stratified analyses

Given that *TLR7* is located on the X chromosome, we performed sex-stratified analyses of the severe disease phenotype to determine if the effect was also observed in females. These could only be done for the M3 and M4 masks due to very low number of M1 mask qualifying variants (**Figure 5**). In both we still see a clear effect among males with a 4.81-fold increase in the odds of severe COVID-19 in M3-variant carriers (95% CI: 2.41-9.59, 5 case carriers, 47 control carriers), and a 3.08-fold increase in M4-variant carriers (95% CI: 1.83-5.20, 7 case carriers, 143 control carriers). In females, we still observed a nominally significant signal in the M3 mask, with a 15.2-fold in odds of severe disease in M3-variant carriers (95% CI: 1.51-153.4). However, an M3-variant was observed in only one female with severe disease (heterozygous) in these analyses (compared to 76 heterozygous controls). In M4 variant, the analyses included 2 female heterozygous carriers (and 203 heterozygous controls), with a 4.86-fold in odds of severe disease (95% CI: 0.43-54.3).

**Figure 5:**
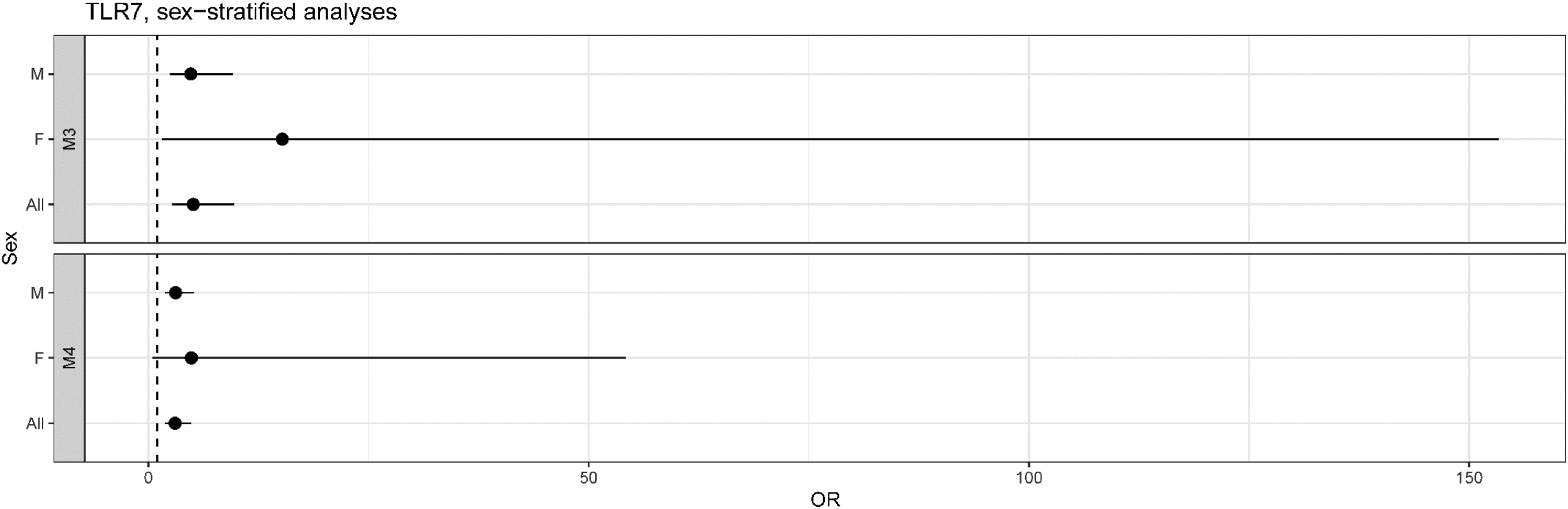
Sex-stratified *TLR7* analyses.

### Rare variants in interferon-related genes and at previously reported genome-wide significant loci

Despite a 7.7-fold increase in number of cases, and a 1,069-fold increase in number of controls, the previously reported associations of genes in the interferon pathway with COVID-19 outcomes^15,16^ could not be replicated with either our exome-wide significance threshold (**Supp. Table 11**) or a more liberal one of p=0.05/10=0.005 (based on Bonferroni correction by the number of genes in the interferon pathway defined in a previous study^15^).

We also tested for rare variant associations between GWAS candidate genes from genome-wide significant loci in the COVID-19 HGI GWAS meta-analyses, but observed no exome-wide significant associations (**Supp. Table 12**). However, at a more liberal Bonferroni threshold of p=0.05/46=0.001 (correcting for the 46 genes in the COVID-19 HGI GWAS associated loci), we observed an increased burden of pLoF (M1) or missense variants (M3 mask) in *ABO* gene among those susceptible to SARS-CoV-2 infection. For example, individuals carrying a pLoF (M1) with MAF<0.1% in *ABO* were at a 2.34-fold higher risk of having a positive SARS-CoV-2 infection (95% CI: 1.50-3.64, p=1.6×10^−4^). The *ABO* results were driven mainly by European and African ancestry participants (**Supp. Figure 13**). Note that deleterious variants in *ABO* often lead to blood groups A and B^36,37^, which is consistent with the epidemiological association that non-type-O individuals are at higher risk of COVID-19^38^. However, more work is required to better understand the genetics of this locus as it relates to COVID-19 outcomes. Lastly, missense variants in *NSF* (mask M4, MAF<1%) were also associated with higher susceptibility to SARS-CoV-2 (OR: 1.48, 95% CI: 1.21-1.82, p=1.4×10^−4^), but this association was not present in other masks (**Supp. Table 12**).

### Replication in GenOMICC

Data for the M1 mask for *TLR7* and *MARK1* in the severe COVID-19 phenotype was then replicated with the GenOMICC cohort^11^, a prospective study enrolling critically ill individuals with COVID-19, with controls selected from the 100,000 genomes cohort^39^. Results are shown in **Table 2**. For *TLR7*, European ancestry individuals with a pLoF (M1) had a 4.70-fold increase in odds of severe disease (95% CI: 1.58 to 14.0, p=0.005). In the sample of South Asian ancestry individuals, a pLoF (M1) was associated with a 1.90-fold increase in odds of severe disease, but the 95% confidence interval crossed the null (0.23 to 15.6, p=0.55), which was likely due to a much smaller sample size than in the European ancestry subgroup (1,202 vs 10,645). Of interest, in both Europeans and South Asians, no pLoFs were observed in either of the control groups.

**Table 2:**
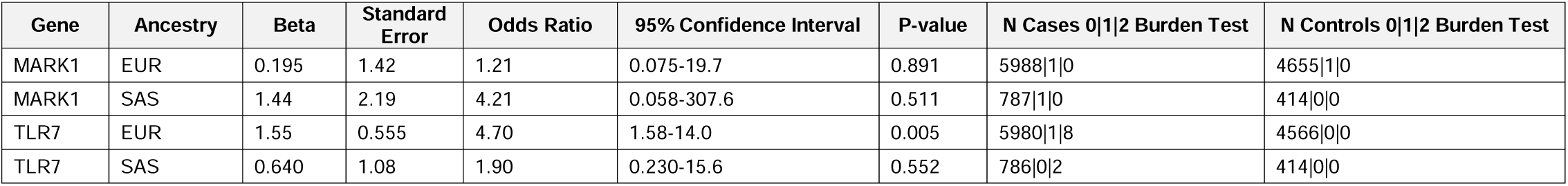
Replication of M1 mask, severe COVID-19, *MARK1* and *TLR7* results in the GenOMICC cohort. Note that the same variants were included in both the MAF<1% and MAF<0.1% replication, and the same results were obtained (shown here).

| Gene | Ancestry | Beta | Standard Error | Odds Ratio | 95% Confidence Interval | P-value | N Cases 0\|1\|2 Burden Test | N Controls 0\|1\|2 Burden Test |
| --- | --- | --- | --- | --- | --- | --- | --- | --- |
| MARK1 | EUR | 0.195 | 1.42 | 1.21 | 0.075-19.7 | 0.891 | 5988\|1\|0 | 4655\|1\|0 |
| MARK1 | SAS | 1.44 | 2.19 | 4.21 | 0.058-307.6 | 0.511 | 787\|1\|0 | 414\|0\|0 |
| TLR7 | EUR | 1.55 | 0.555 | 4.70 | 1.58-14.0 | 0.005 | 5980\|1\|8 | 4566\|0\|0 |
| TLR7 | SAS | 0.640 | 1.08 | 1.90 | 0.230-15.6 | 0.552 | 786\|0\|2 | 414\|0\|0 |

**Table 3:** Results of burden tests at genes identified from common variants GWAS in the COVID-19 HGI. Only genes with p<0.05/46 are shown here. Full results available in **Supp. Table 11**.

| Gene | Mask | Phenotype | MAF | Beta | Standard Error | Odds Ratio | 95% Confidence Interval | P-value | Heterogeneity p-value | N Cases 0\|1\|2 Burden Test | N Controls 0\|1\|2 Burden Test |
| --- | --- | --- | --- | --- | --- | --- | --- | --- | --- | --- | --- |
| NSF | M4 | Susceptibility | <1% | 0.395 | 0.104 | 1.484 | 1.21-1.82 | $1.44 \times 10^{-4}$ | 0.866 | 25752\|127\|2 | 585642\|1907\|5 |
| ABO | M1 | Susceptibility | <0.1% | 0.851 | 0.226 | 2.341 | 1.50-3.65 | $1.68 \times 10^{-4}$ | 0.498 | 22778\|27\|0 | 572310\|296\|0 |
| ABO | M1 | Susceptibility | <1% | 0.784 | 0.209 | 2.19 | 1.45-3.30 | $1.75 \times 10^{-4}$ | 0.826 | 23460\|34\|0 | 574608\|364\|0 |
| ABO | M3 | Susceptibility | <1% | 0.729 | 0.195 | 2.073 | 1.41-3.04 | $1.89 \times 10^{-4}$ | 0.869 | 24455\|42\|0 | 575051\|434\|0 |
| ABO | M1 | Hospitalisation | <0.1% | 1.33 | 0.395 | 3.78 | 1.74-8.20 | $7.56 \times 10^{-4}$ | 0.542 | 7859\|12\|0 | 561642\|291\|0 |
| ABO | M1 | Susceptibility | <0.1% | 0.736 | 0.222 | 2.088 | 1.35-3.23 | $9.35 \times 10^{-4}$ | 0.512 | 23779\|29\|0 | 572799\|320\|0 |

On the other hand, we could not replicate an effect from *MARK1*, which demonstrated an OR of 1.21 in European ancestry participants (95% CI 0.075 to 19.7, p=0.89) and an OR of 4.21 in South Asian ancestry individuals (95% CI 0.058 to 307, p=0.51).

## Discussion

Whole genome and whole exome sequencing can provide unique insights into genetic determinants of COVID-19, by uncovering associations between rare genetic variants and COVID-19. Specifically, gene burden tests can be particularly helpful, because they test for coding variants, thereby pointing directly to a causal gene and often suggesting a direction of effect. However, such studies require careful control for population stratification and an adapted analysis method such as burden testing, in order to have enough statistical power to find those associations. In our study, we observed that individuals with rare deleterious variants at *TLR7* are at increased risk of severe COVID-19 (up to 13.1-fold increase in odds in those with pLoFs). Although this association was suggested by previous studies^28–30^, our study provides the most definitive evidence for the role of TLR7 in COVID-19 pathogenesis, with exome-wide significance for this gene in the discovery phase followed by strong replication in a large independent cohort. *TLR7* is a well-studied part of the antiviral immunity cascade and stimulates the interferon pathway after recognizing viral pathogen-associated molecular patterns. Given its location on the X chromosome, it has been hypothesis that it could partly explain the observed COVID-19 outcome differences between sexes^40–42^, and to our knowledge, this is the first study to show that even in heterozygous females, this gene can potentially play a role in severe disease. Further, this our results suggest that *TLR7* mediated genetic predisposition to severe COVID-19 may be a dominant or co-dominant trait, an observation that cannot be made in cohorts limited to male participants^28,30^.

We also uncovered a potential role for cellular microtubule disruption in the pathogenesis of COVID-19 and the microtubule network is known to be exploited by other viruses during infections^43^. Indeed, the MARK1 protein has been shown to interact with SARS-CoV-2 in previous *in-vitro* experiments^33^. Nevertheless, these findings at *MARK1* were not replicated in the GenOMICC cohort and will need to be tested in larger cohorts, especially given the small number of highly deleterious variants that we found in our consortium. Lastly, we found single variant associations at *IL6R, SRRM1*, and *FRMD5*. While *IL6R* is is already a therapeutic target^44,45^ for COVID-19, and *SRRM1* has been reported in a previous pre-print^46^, these were found in smaller cohorts and will require replication.

To our knowledge, this is the first time a rare variant burden test meta-analysis has been attempted on such a large scale. Our framework allowed for easy and interpretable summary statistics results, while at the same time preventing participant de-identification or any breach of confidentiality that stems from sharing results of rare genetic variant analyses^47^. It also provides important insights into how these endeavours should be planned in the future. First, our burden test operated under the assumption that the effect of any of the deleterious variants on the phenotype would be in the same direction and did not account for compound deleterious variant heterozygosity. This allowed for easier meta-analysis across cohorts, but may have decreased statistical power. Other methods may be needed in future analysis to soften this assumption, though some of these cannot be easily meta-analyzed across multiple cohorts directly from summary statistics (e.g., SKAT-O^48^). Similarly, methods that combine both rare and common variants might also provide additional insights into disease outcomes^31,49^. Second, our results highlight the importance at looking at different categories of variants through different masks to increase sensitivity and specificity of our burden tests. Third, while the largest biobanks contributed the most to the signal observed at *TLR7* and *MARK1*, many of our smaller prospective COVID-19 specific cohorts also contributed to the signal. This further highlights the importance of robust study design to improve statistical power, especially with rare variant associations. Lastly, work remains to be done to standardize sequencing and annotation pipelines to allow comparisons of results easily across studies and cohorts. Here, we provided a pipeline framework to every participating cohort, but there remains room for process harmonization. While the decentralized approach to genetic sequencing, quality control, and analyses allowed for more rapid generation of results, it may come at the cost of larger variance in our estimates. In the future, more sophisticated approaches may be required to increase statistical power of exome-wide rare variant association studies^50^.

Our study had limitations. First, even if this is one of the world’s largest consortia using sequencing technologies for the study of rare variants, we remain limited by a relatively small sample size. For example, in a recent analyses of UK Biobank exomes, many of the phenotypes for which multiple genes were found using burden tests had a much higher number of cases than in our analyses (e.g. blonde hair colour, with 48,595 cases)^22^. Further, rare variant signals were commonly found in regions enriched in common variants found in GWASs. The fact that *ABO* and *NSF* were the only genes from the COVID-19 HGI GWAS that were also identified in our burden test (albeit using a more liberal significance threshold), also suggests a lack of statistical power. Similarly, GenOMICC, a cohort of similar size, was also unable to find rare variant associations using burden tests^11^. However, their analysis methods were different from ours, making further comparisons difficult. Nevertheless, this provides clear guidance that smaller studies looking at the effect of rare variants across the genome are at considerable risk of finding both false positive and false negative associations. Second, many cohorts used population controls, which may have decreased statistical power given that some controls may have been misclassified. However, given that COVID-19 critical illness remains a rare phenomenon^51^, our severe disease phenotype results are unlikely to be strongly affected by this. Finally, the use of population control is a long-established strategy in GWAS burden tests^7,8,11,22,52^, and the statistical power gain from increasing our sample size is likely to have counter-balanced the misclassification bias.

In summary, we reproduced an exome-wide significant association with severe COVID-19 outcomes in carriers of rare deleterious variants at *TLR7*, for both sexes. Our results also suggest an association between the cellular microtubule network and severe disease, which requires further validation. More importantly, our results underline the fact that future genome-wide studies of rare variants will require considerably larger sample size, but our work provides a roadmap for such collaborative efforts.

## Methods

### COVID-19 outcome phenotypes

For all analyses, we used three case-control definitions: A) Severe COVID-19, where cases were those who died, or required either mechanical ventilation (including extracorporeal membrane oxygenation), high-flow oxygen supplementation, new continuous positive airway pressure ventilation, or new bilevel positive airway pressure ventilation, B) Hospitalized COVID-19, where cases were all those who died or were admitted with COVID-19, and C) Susceptibility to COVID-19, where cases are anyone who tested positive for COVID-19, self-reported an infection to SARS-CoV-2, or had a mention of COVID-19 in their medical record. For all three, controls were individuals who did not match case definitions, including population controls for which case status was unknown (given that most patients are neither admitted with COVID-19, nor develop severe disease^53^). These three analyses are also referred to as analyses A2, B2, and C2 by the COVID-19 Host Genetics Initiative^8^, respectively.

### Cohort inclusion criteria and genetic sequencing

Any cohort with access to genetic sequencing data and the associated patient level phenotypes were allowed in this study. Specifically, both whole-genome and whole-exome sequencing was allowed, and there were no limitations in the platform used. There were no minimal number of cases or controls necessary for inclusion. However, the first step of Regenie, which was used to perform all tests (see below), uses a polygenic risk score which implicitly requires that a certain sample size threshold be reached (which depends on the phenotype and the observed genetic variation). Hence, cohorts were included if they were able to perform this step. All cohorts obtained approval from their respective institutional review boards, and informed consent was obtained from all participants. More details on each cohort’s study design and ethics approval can be found in the **Supp Tables 1-2**.

### Variant calling and quality control

Variant calling was performed locally by each cohort, with the pre-requisite that variants should not be joint-called separately between cases and controls. Quality control was also performed individually by each cohort according to individual needs. However, a general quality control framework was made available using the Hail software^54^. This included variant normalization and left alignment to a reference genome, removal of samples with call rate less than 97% or mean depth less than 20. Genotypes were set to unknown if they had genotype quality less than 20, depth less than 10, or poor allele balance (more than 0.1 for homozygous reference calls, less than 0.9 for homozygous alternative calls, and either below 0.25 or above 0.75 for heterozygous calls. Finally, variants were removed from if the mean genotype quality was less than 11, mean depth was less than 6, mean call rate less than or equal to 0.8, and Hardy-Weinberg equilibrium p-value less than or equal to 5×10^−8^ (10^−16^ for single variant association tests). Details on variant calling and quality control is described for each cohort in the **Supp. Table 1**.

### Single variant association tests

We performed single variant association tests using a GWAS additive model framework, with the following covariates: age, age^2^, sex, age*sex, age^2^*sex, 10 genetic principal components obtained from common genetic variants (MAF>1%). Each cohort performed their analyses separately for each genetic ancestry, but also restricted their variants to those with MAF>0.1% and MAC>6. Summary statistics were then meta-analyzed using a fixed effect model within each ancestry and using a DerSimonian-Laird random effect model across ancestries with the Metal package^55^ and its random effect extension^56^. Lastly, given that multiple technologies were used for sequencing, and that whole-exome sequencing can provide variant calls of worse quality in its off-target regions^57^, we used the UKB, GHS, and Penn Medicine whole-exome sequencing variants as our “reference panel” for whole-exome sequencing. Hence, only variants reported in at least one of these biobanks were used in the final single-variant analyses.

### Variant exclusion list

For the burden tests, we also compiled a list of variants that had a MAF > 1 % or > 0.1 % in any of the participating cohorts. This list was used to filter out variants that were less likely to have a true deleterious effect on COVID-19, even if they were considered rare in other cohorts, or in reference panels^25^. We created two such variant exclusion lists: one to be used in our burden test with variants of MAF less than 1%, and the other for the analysis with MAF less than 0.1%. In any cohort, if a variant had a minor allele count of 6 or more, and a MAF of more than 1% (or 0.1%), this variant was added to our exclusion list. This list was then shared with all participating cohorts, and all variants contained were removed from our burden tests.

### Gene burden tests

The following analyses generally followed the methods used by recent literature on large-scale whole-exome sequencing^22^ and the COVID-19 HGI^8^.

The burden tests were performed by pooling variants in three different variant sets (called masks), as described in recent UK Biobank whole-exome sequencing papers by Backman *et al*.^22^ and Kosmicki *et al*.^17^. : “M1” which included loss of functions as defined by high impact variants in the Ensembl database^23^ (i.e. transcript ablation, splice acceptor variant, splice donor variant, stop gained, frameshift variant, stop lost, start lost, transcript amplification), “M3” which included all variants in M1 as well as moderate impact indels and any missense variants that was predicted to be deleterious based on all of the *in-silico* pathogenicity prediction scores used, and “M4” which included all variants in M3 as well as all missense variants that were predicted to be deleterious in at least one of the *in-silico* pathogenicity prediction scores used. For *in-silico* prediction, we used the following five tools: SIFT^58^, LRT^59^, MutationTaster^60^, PolyPhen2^61^ with the HDIV database, and PolyPhen2 with the HVAR database. Protein coding variants were collapsed on canonical gene transcripts.

Once variants were collapsed into genes in each participant, for each mask, genes were given a score of 0 if the participant had no variants in the mask, a score of 1 if the participant had one or more heterozygous variant in this mask, and a score of 2 if the participant had one or more homozygous variant in this mask. These scores were used as regressors in logistic regression models for the three COVID-19 outcomes above. These regressions were also adjusted for age, age^2^, sex, age*sex, age^2^*sex, 10 genetic principal components obtained from common genetic variants (MAF>1%), and 20 genetic principal components obtained from rare genetic variants (MAF<1%). The Regenie software^18^ was used to perform all burden tests, and generate the scores above. Regenie uses Firth penalized likelihood to adjust for rare or unbalanced events, providing unbiased effect estimates.

All analyses were performed separately for each of six genetic ancestries (African, Admixed American, East Asian, European, Middle Eastern, and South Asian). Summary statistics were meta-analyzed as for the single variant analysis. Participant assignment to genetic ancestry was done locally by each cohort, more details on the methods can be found in the **Supp. Table 1**.

Lastly, we used ACAT^35^ to meta-analyze p-values across masks, within each phenotype separately. ACAT is not affected by lack of independence between tests. These values were used to draw Manhattan and QQ plots in **Figure 2**.

## Code availability

Code guidance is available at https://github.com/DrGBL/WES.WGS.

## Data availability

The exome-wide burden test summary statistics are available in the Supplements. The single variant association studies summary statistics will be made available openly on the GWAS Catalog^62^ shortly after publication.

## Data Availability

The exome-wide burden test summary statistics are available in the Supplements. The single variant association studies summary statistics will be made available openly on the GWAS Catalog following publication.

## Supplements

**Supplementary Table**: Contains **Supp. Tables 1-12**.

*Supplementary Table 1*: cohorts demographics and sequencing details.

*Supplementary Table 2*: acknowledgement and conflicts of interests

*Supplementary Table 3*: Genentech co-authors

*Supplementary Table 4*: DeCOI co-authors

*Supplementary Table 5*: GEN-COVID (Italy) co-authors

*Supplementary Table 6*: GEN-COVID (Spain) co-authors

*Supplementary Table 7*: Mount Sinai Biobank co-authors

*Supplementary Table 8*: Inflation factors

*Supplementary Table 9*: Single variant association tests significant results

*Supplementary Table 10*: Full Burden test results

*Supplementary Table 11*: IFN related genes burden test results

*Supplementary Table 12*: GWAS loci burden test results

**Supplementary Figures**: Contains **Supp. Figures 1-12**.

*Supplementary Figure 1*: QQ plot and Manhattan plot for the exome-wide single variant association studies.

*Supplementary Figure 2*: chromosome 3 single variant association studies result by cohort for the severe disease phenotype.

*Supplementary Figure 3*: chromosome 3 single variant association studies result by cohort for the hospitalized disease phenotype.

*Supplementary Figure 4*: chromosome 3 single variant association studies result by cohort for the susceptibility disease phenotype.

*Supplementary Figure 5*: Single variant association study results at the three novel loci.

*Supplementary Figure 6*: QQ plot from exome burden test ACAT meta-analyses.

*Supplementary Figure 7*: *TLR7* ancestry and cohort specific results for MAF<0.1% and severe phenotype

*Supplementary Figure 8*: *TLR7* ancestry and cohort specific results for MAF<1% and severe phenotype

*Supplementary Figure 9*: *MARK1* ancestry and cohort specific results for MAF<0.1% and severe phenotype

*Supplementary Figure 10*: *MARK1* ancestry and cohort specific results for MAF<1% and severe phenotype

*Supplementary Figure 11*: *MARK1* ancestry and cohort specific results for MAF<0.1% and hospitalized phenotype

*Supplementary Figure 12*: *MARK1* ancestry and cohort specific results for MAF<1% and hospitalized phenotype

*Supplementary Figure 13*: ancestry stratified results for *TLR7, MARK*, and *ABO* burden tests.

## Acknowledgement

We thank the patients who volunteered to all participating cohorts, and the researchers and clinicians who enrolled them into the respective studies. A full list of acknowledgements can be found in **Supp. Table 2-7**.

## Author contributions

Conceptualization and methodology: GBL, GP, JAK, ETC, TD, SF, CS, ASchmidt, PO, MQ, EÇ, KKundu, KV, TNakanishi, MSAbedalthagafi, HZ, SOssoswksi, ECS, KUL, HM, DBG, KKiryluk, AR, MARF, JBR

Formal analyses: GBL, GP, JAK, ETC, TD, SF, CS, ASchmidt, PO, MQ, EÇ, KKundu, KW, JJ, ADS, LGS, BPrzychodzen, TC, TDP, AStuckey, AS, YK, YO, AR

Investigation: GBL, GP, JAK, ETC, TD, SF, CS, ASchmidt, PO, UK, MQ, EÇ, KKundu, KW, JJ, ADS, LGS, DMJ, RCT, DDV, NSimons, EC, RS, EES, SSK, SG, MMerad, JDB, NDB, AWC, BPrzychodzen, TC, TDP, NShang, FB, FF, FM, KC, MN, SP, JKB, AStuckey, ASalas, XB, JPS, AGC, IRT, FMT, AGanna, KJK, KV, MB, GB, RJME, VF, DM, DL, MLathrop, VM, TNakanishi, RF, MH, MLipcsey, YMZ, JN, KMSB, WL, Abolze, SW, SR, FT, ES, IN, SD, Ncasadei, SMotameny, MA, SMassadeh, NA, MSAlmutairi, YMA, SAA, FSAH, AA, FA, Salotaibi, Abinowayn, EAA, HEB, MF, DHG, SArteaga, AStephens, MJB, PCB, TNY, ST, SE, TS, PJT, MEB, YP, AGonzalez, Nchavan, RJ, BPasaniuc, BY, SS, CR, SB, PYB, MD, PZ, MSypniewski, EK, PC, VN, NH, VS, MP, CP, WC, KT, MSugiyama, YK, TH, TNaito, HN, RE, AK, SOgawa, TK, KF, YO, SI, SMiyano, SManghul, MSAbedalthagafi, HZ, JJG, NLW, SOssoswksi, KUL, ECS, OR, MMoniuszko, MK, HM, SII, AV, DBG, KKiryluk, AR, MARF, JBRResources: ETC, KW, AWC, FM, JKB, MB, GB, RJME, VF, DM, MLathrop, VM, MH, RF, MLipcsey, BPasaniuc, BY, SS, CR, SB, PYB, EK, PL, YK, YO, SManghul, MSAbedalthagafi, HZ, SOssoswksi, KUL, ECS, OR, MK, HM, SII, AV, DBG, KKiryluk, AR, MARF, JBR

Data curation: GBL, GP, JAK, ETC, TD, SF, CS, ASchmidt, PO, MQ, EÇ, KKundu, KW, JJ, ADS, KUL, LGS, BPrzychodzen, TC, TDP, YK, YO

Writing – original draft: GBL

Writing – review & editing: GBL, GP, JAK, ETC, TD, SF, CS, ASchmidt, PO, UK, MQ, EÇ, KKundu, KW, JJ, ADS, LGS, DMJ, RCT, DDV, NSimons, EC, RS, EES, SSK, SG, MMerad, JDB, NDB, AWC, BPrzychodzen, TC, TDP, NShang, FB, FF, FM, KC, MN, SP, JKB, AStuckey, ASalas, XB, JPS, AGC, IRT, FMT, AGanna, KJK, KV, MB, GB, RJME, VF, DM, DL, MLathrop, VM, TNakanishi, RF, MH, MLipcsey, YMZ, JN, KMSB, WL, Abolze, SW, SR, FT, ES, IN, SD, Ncasadei, SMotameny, MA, SMassadeh, NA, MSAlmutairi, YMA, SAA, FSAH, AA, FA, Salotaibi, Abinowayn, EAA, HEB, MF, DHG, SArteaga, AStephens, MJB, PCB, TNY, ST, SE, TS, PJT, MEB, YP, AGonzalez, Nchavan, RJ, BPasaniuc, BY, SS, CR, SB, PYB, MD, PZ, MSypniewski, EK, PC, VN, NH, VS, MP, CP, WC, KT, MSugiyama, YK, TH, TNaito, HN, RE, AK, SOgawa, TK, KF, YO, SI, SMiyano, SManghul, MSAbedalthagafi, HZ, JJG, NLW, SOssoswksi, KUL, ECS, OR, MMoniuszko, MK, HM, SII, AV, DBG, KKiryluk, AR, MARF, JBR

Visualization: GBL

Supervision: ETC, KW, AWC, FM, JKB, MH, VM, MH, RF, MLipcsey, BPasaniuc, BY, SS, CR, SB, AS, PYB, MS, EK, PL, YK, YO, SManghul, MSAbedalthagafi, HZ, SOssoswksi, KUL, ECS, OR, MK, HM, SII, AV, DBG, KKiryluk, AR, MARF, JBR

Project administration: GBL, FF, FM, AG, DBG, KKiryluk, AR, MARF, JBR

## Materials & Correspondence

J Brent Richards is the corresponding author.

