## Supplementary Figures for "Exome-wide association study to identify rare variants influencing COVID-19 outcomes: Results from the Host Genetics Initiative"

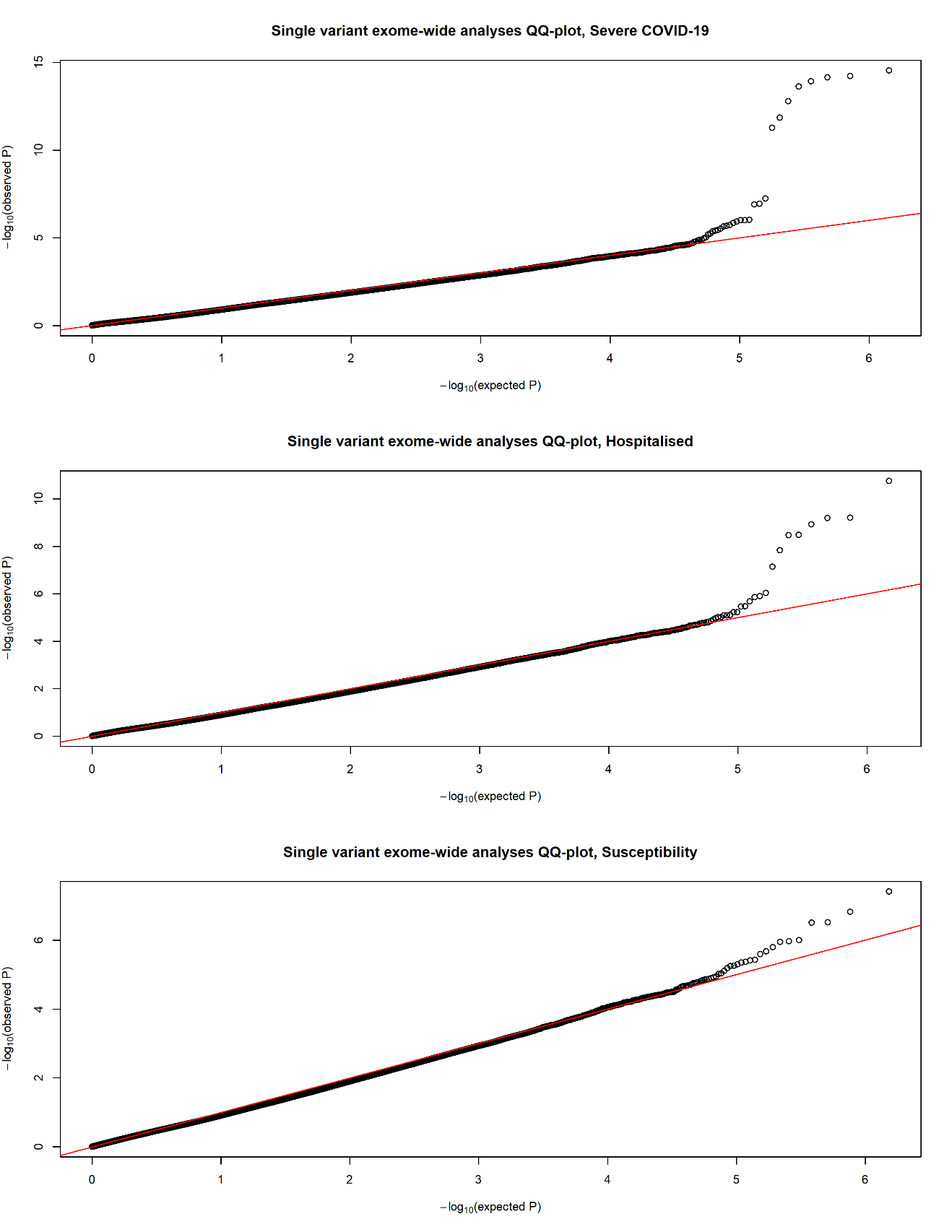

**Supplemental Figure 1**: QQ plot and Manhattan plot for the exome-wide single variant association studies.

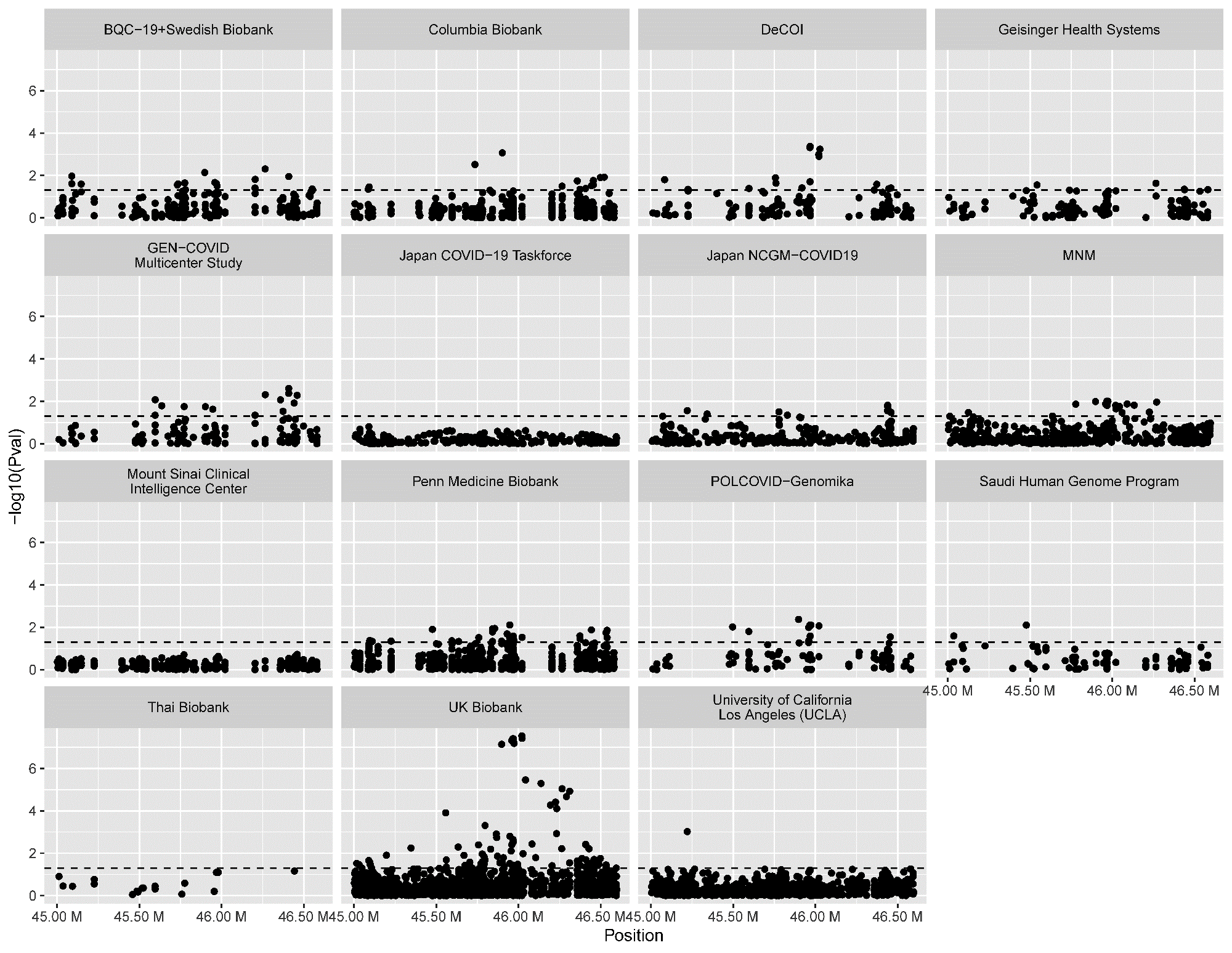

**Supplemental Figure 2**: Chromosome 3 exome single variant association studies result by cohort for the severe disease phenotype. Note that all ancestries are shown together. Black dashed line represents the nominal statistical significance threshold (p=0.05).

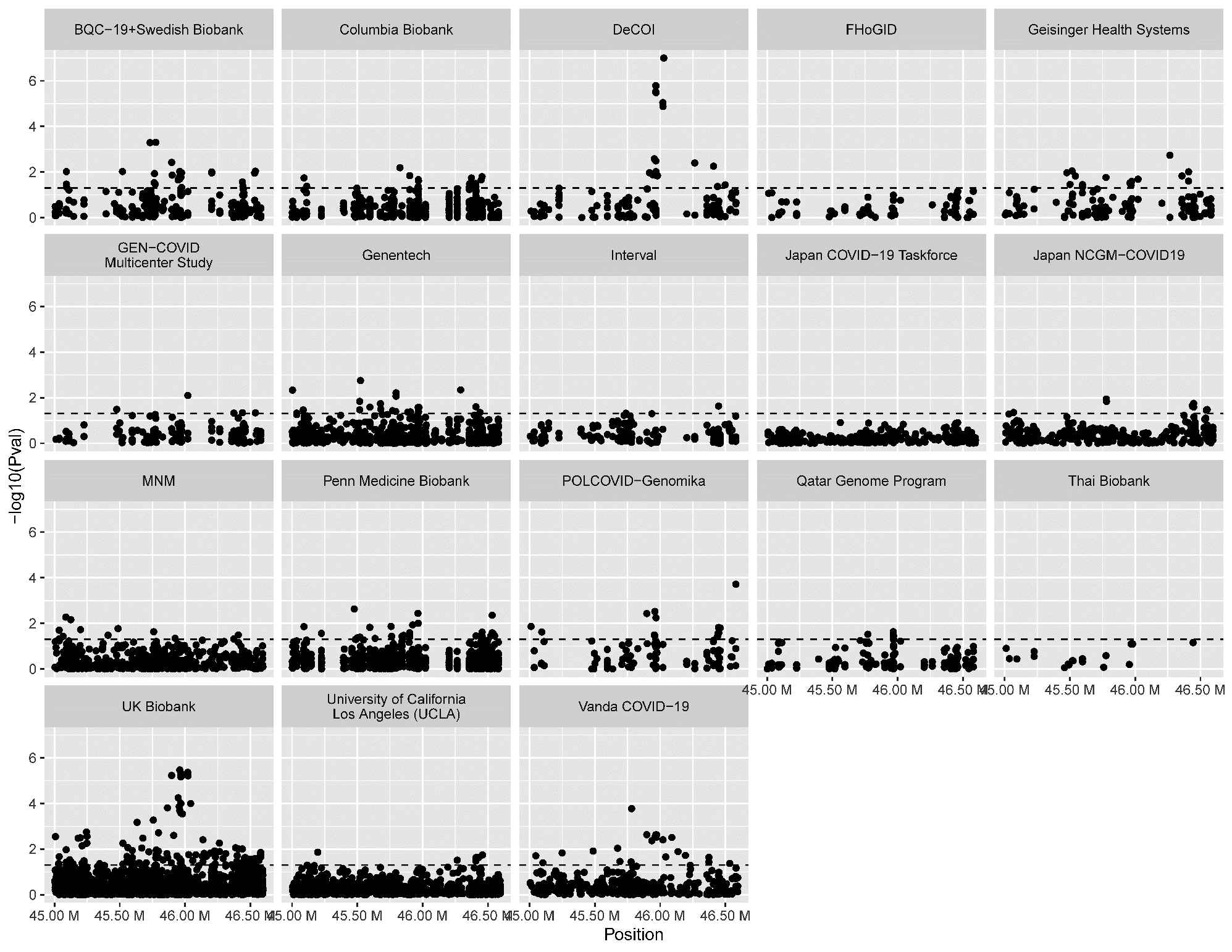

**Supplemental Figure 3**: Chromosome 3 exome single variant association studies result by cohort for the severe disease phenotype. Note that all ancestries are shown together. Black dashed line represents the nominal statistical significance threshold (p=0.05).

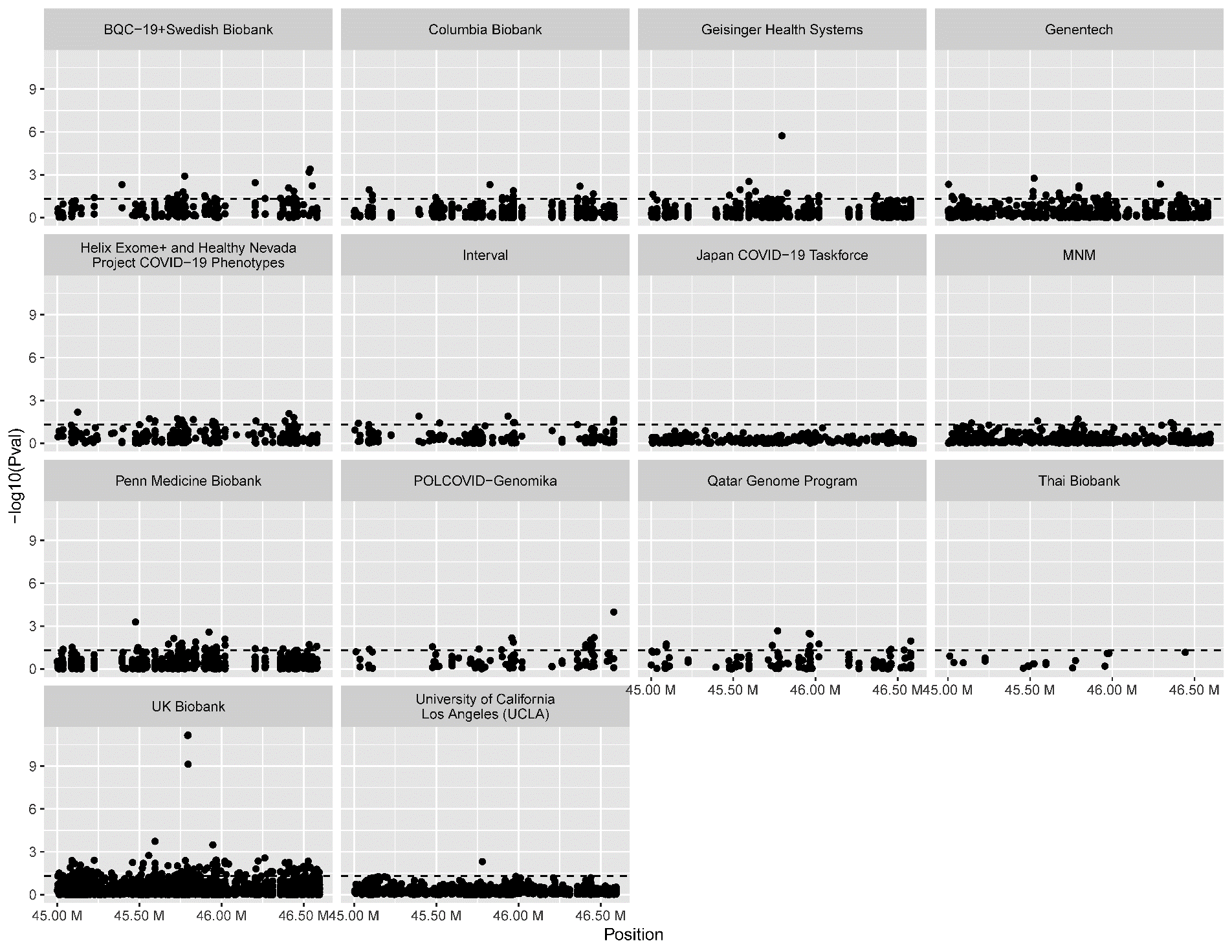

**Supplemental Figure 4**: Chromosome 3 exome single variant association studies result by cohort for the severe disease phenotype. Note that all ancestries are shown together. Black dashed line represents the nominal statistical significance threshold (p=0.05).

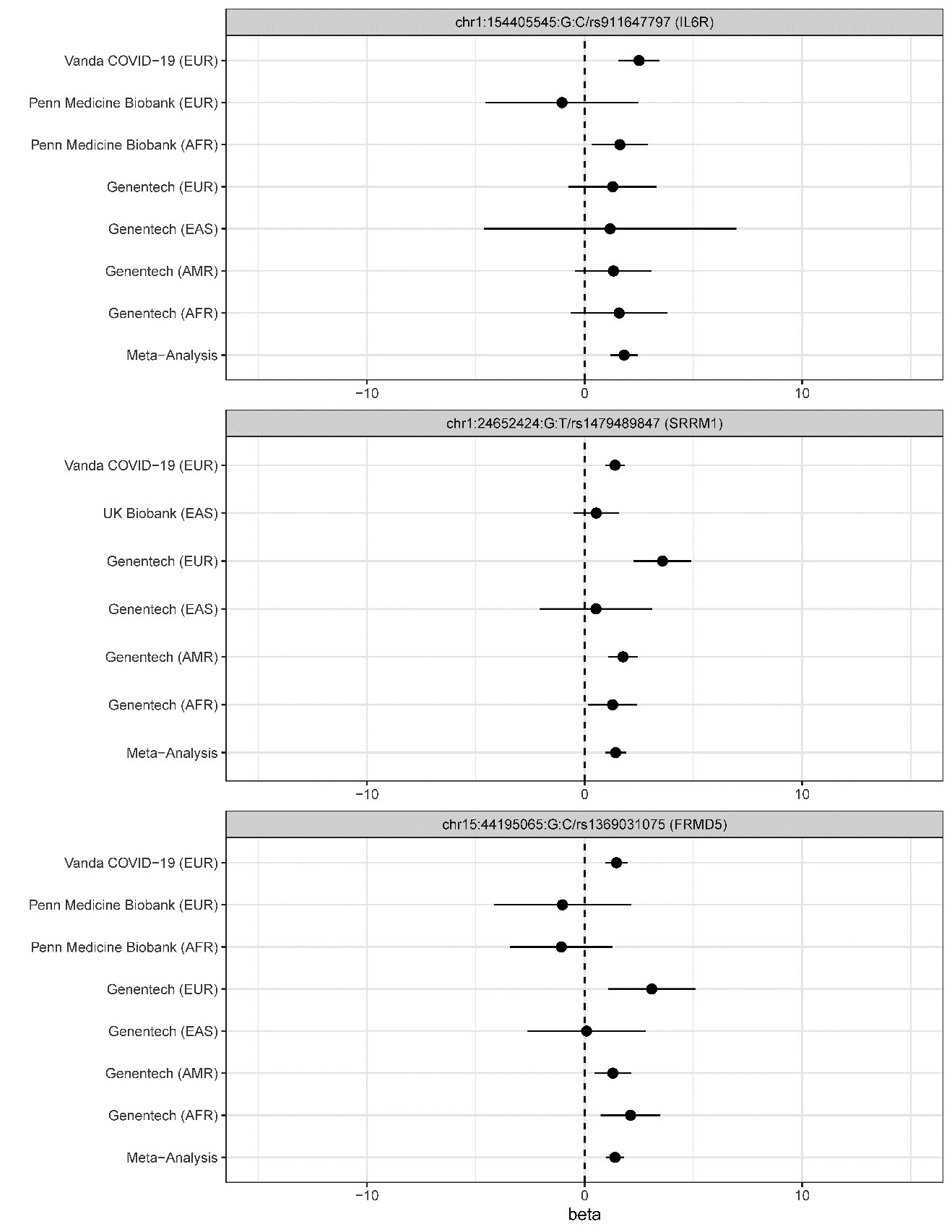
**Supplemental Figure 5**: Single variant association study results at the three novel loci (OR and 95% CI).

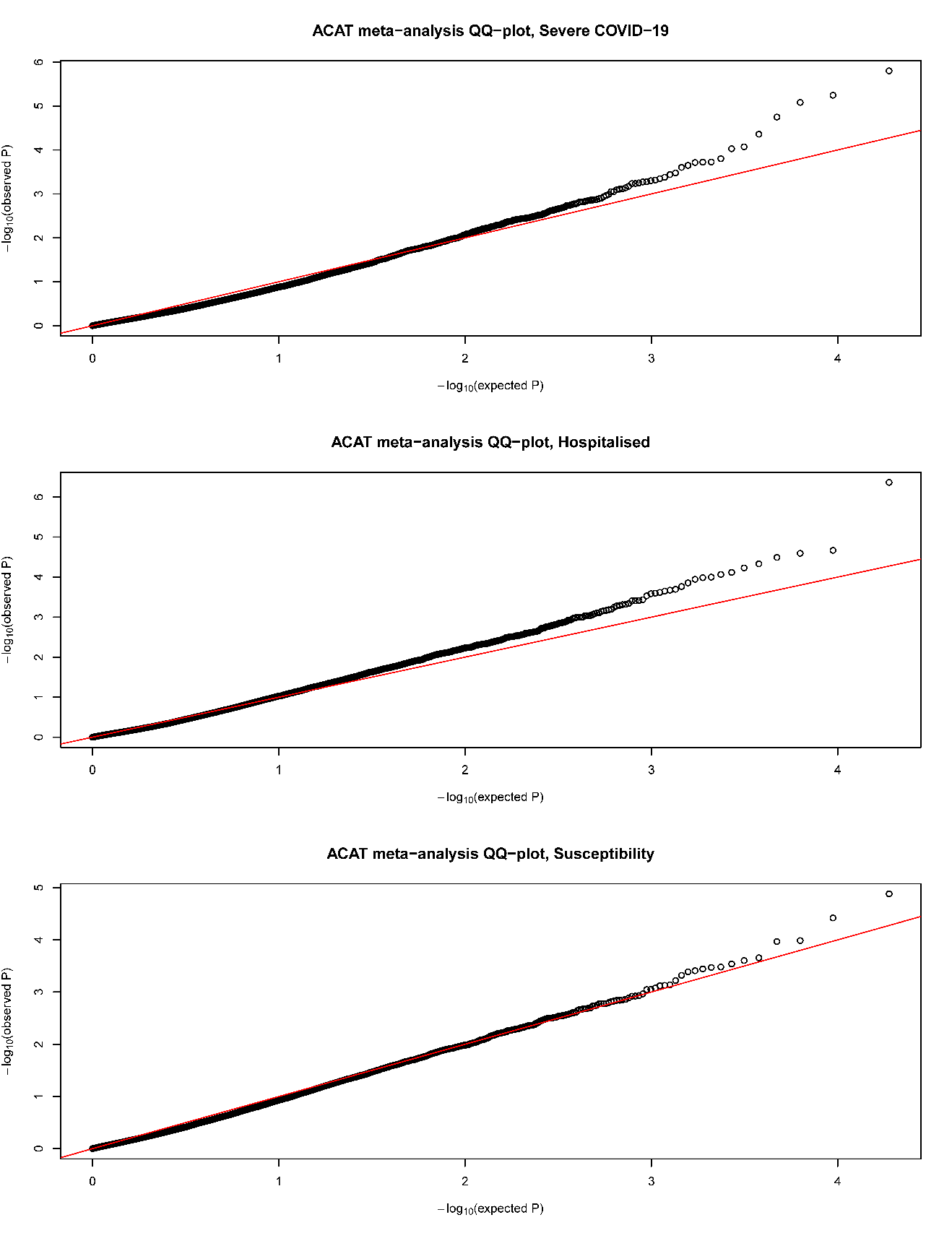

**Supplemental Figure 6**: QQ plot from exome burden test ACAT meta-analyses.

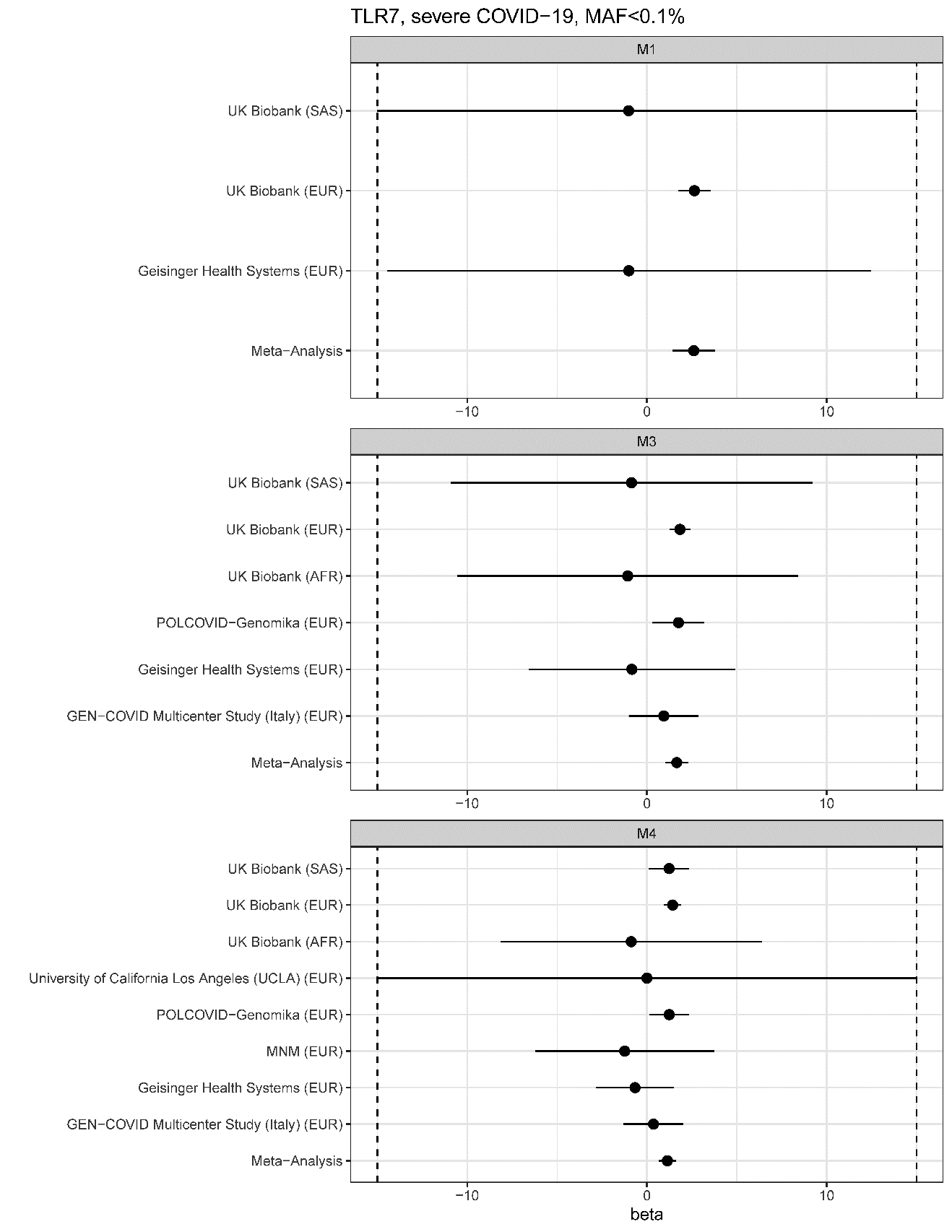

**Supplemental Figure 7**: *TLR7* beta coefficients (on logistic scale) with 95% interval for the severe COVID-19 phenotype, MAF<0.1%. X-axes are cut at -10 and 10.

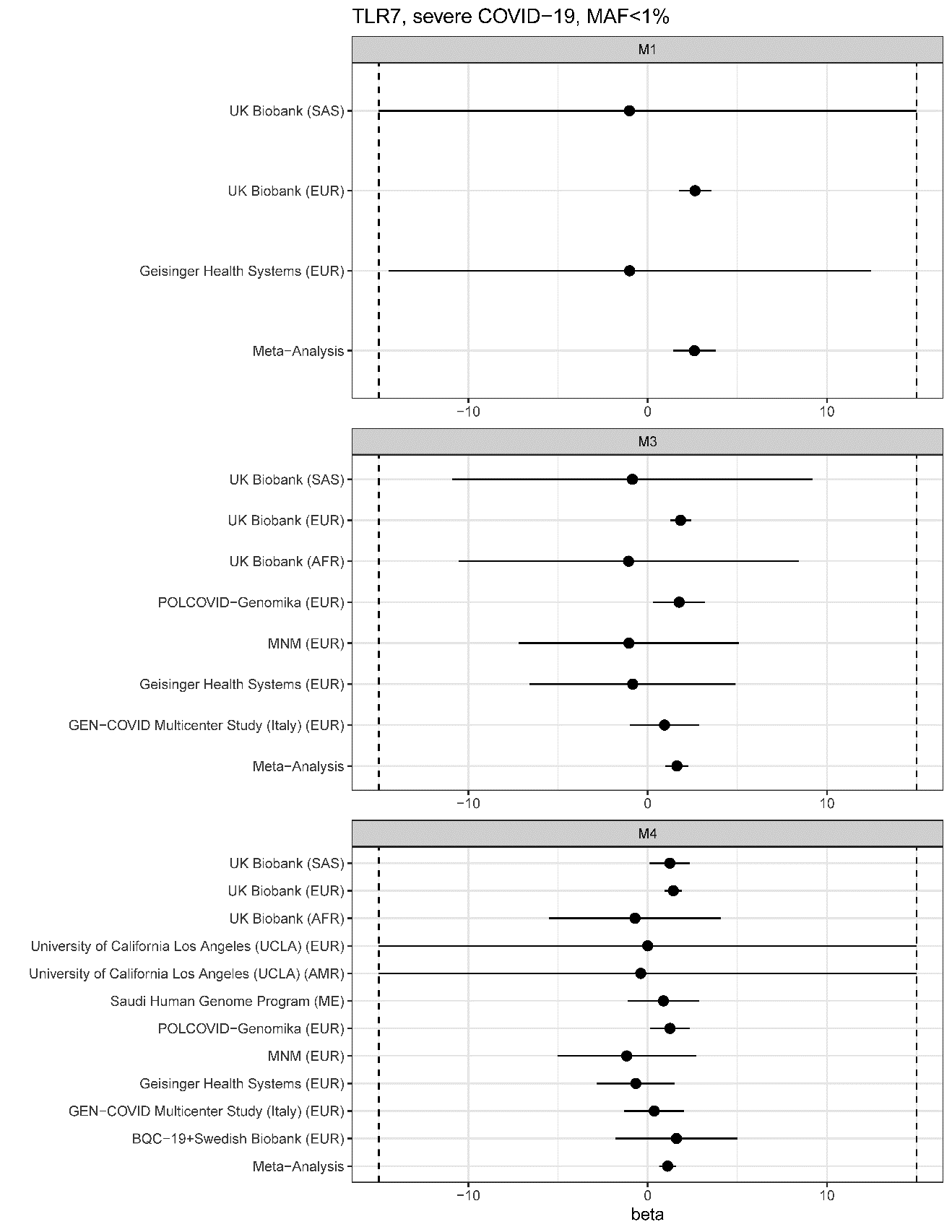

**Supplemental Figure 8**: *TLR7* beta coefficients (on logistic scale) with 95% interval for the severe COVID-19 phenotype, MAF<1%. X-axes are cut at -10 and 10.

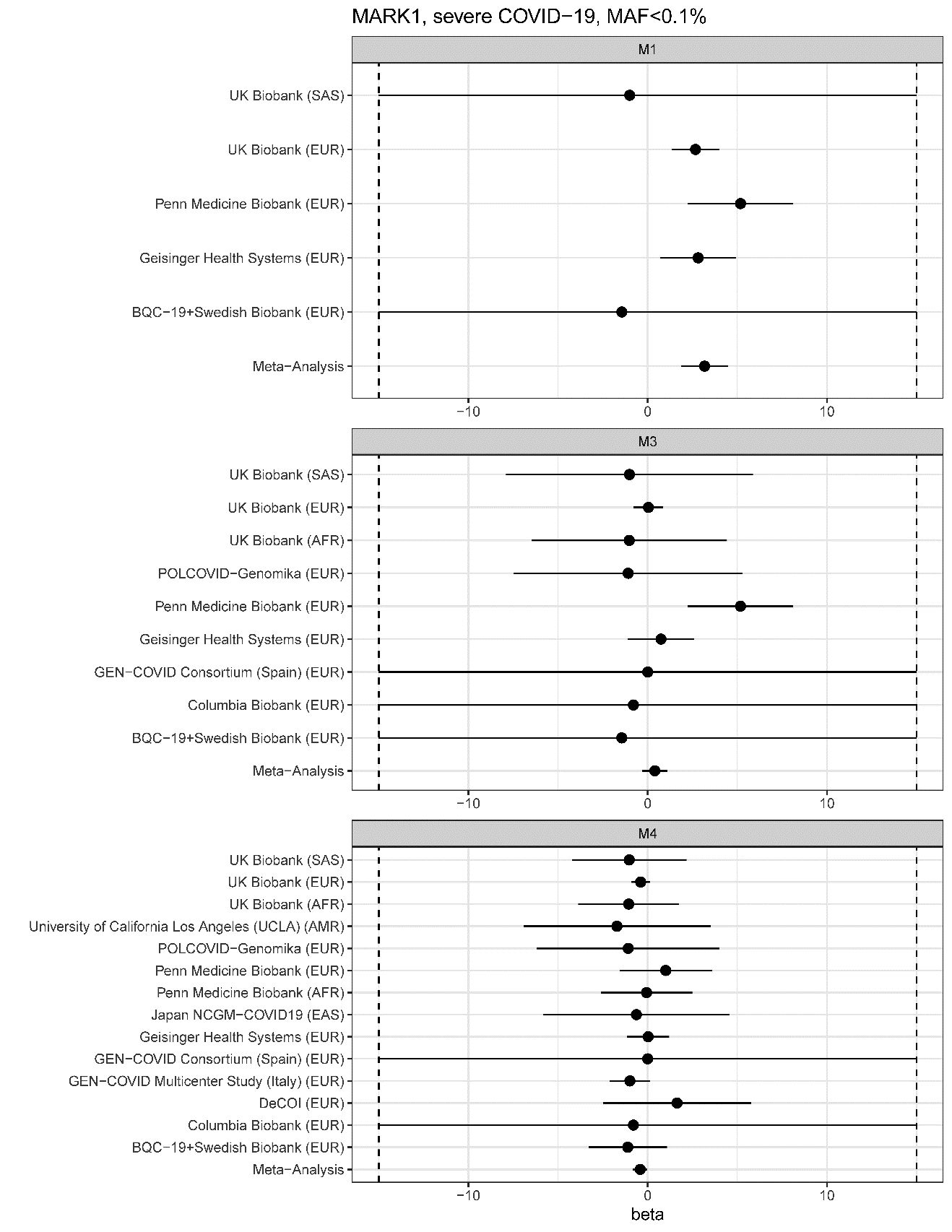

**Supplemental Figure 9**: *MARK1* beta coefficients (on logistic scale) with 95% interval for the severe COVID-19 phenotype, MAF<0.1%. X-axes are cut at -10 and 10.

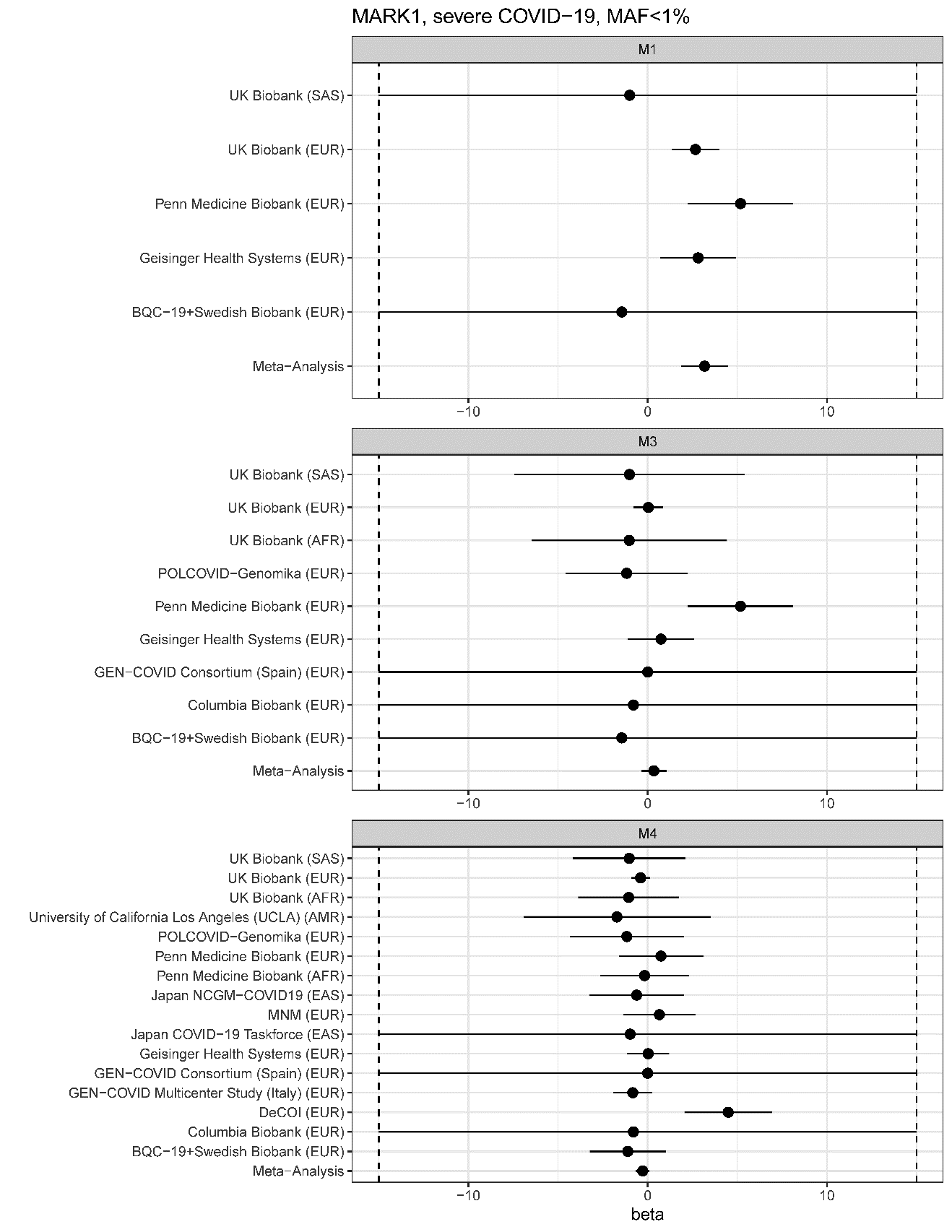

**Supplemental Figure 10**: *MARK1* beta coefficients (on logistic scale) with 95% interval for the severe COVID-19 phenotype, MAF<1%. X-axes are cut at -10 and 10.

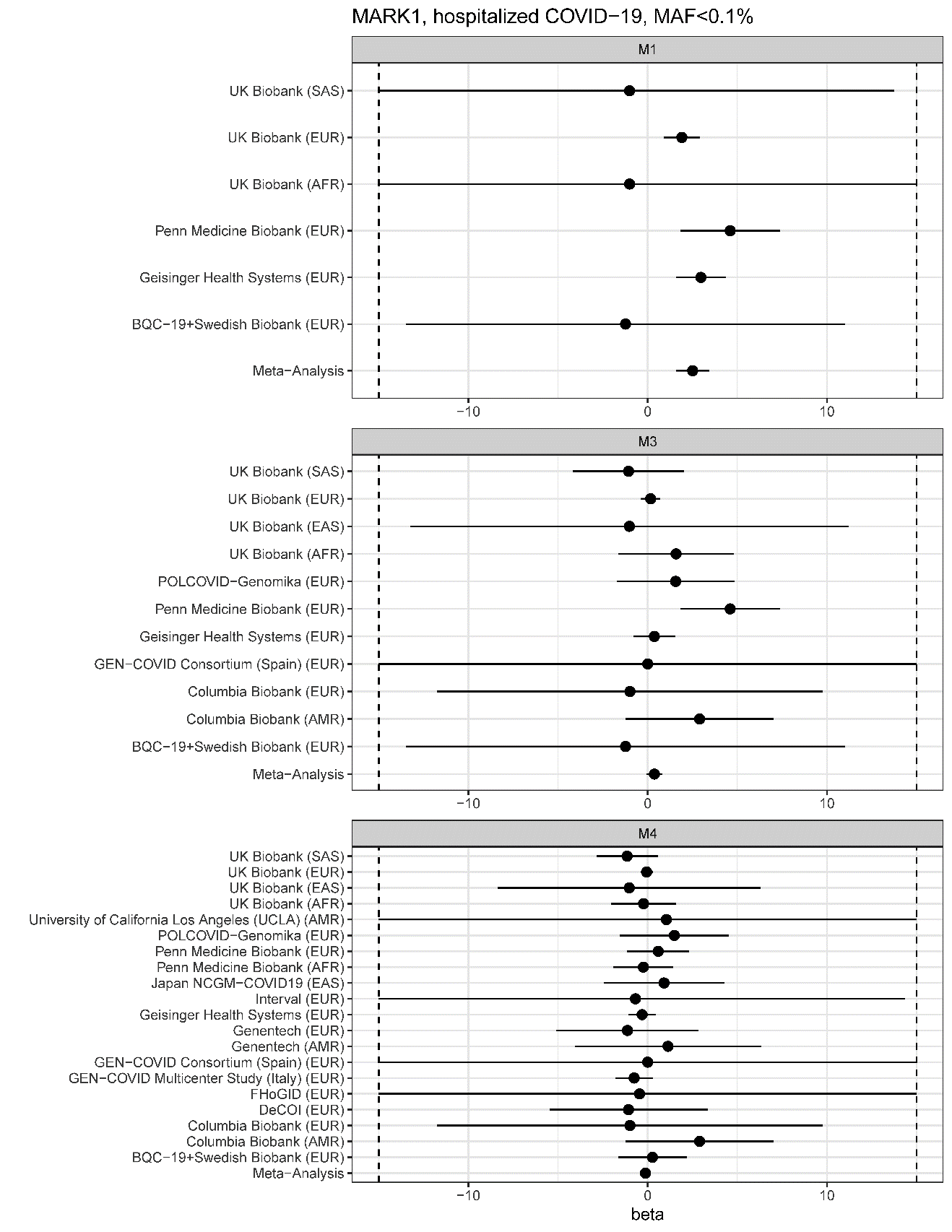

**Supplemental Figure 11**: *MARK1* beta coefficients (on logistic scale) with 95% interval for the hospitalized COVID-19 phenotype, MAF<0.1%. X-axes are cut at -10 and 10.

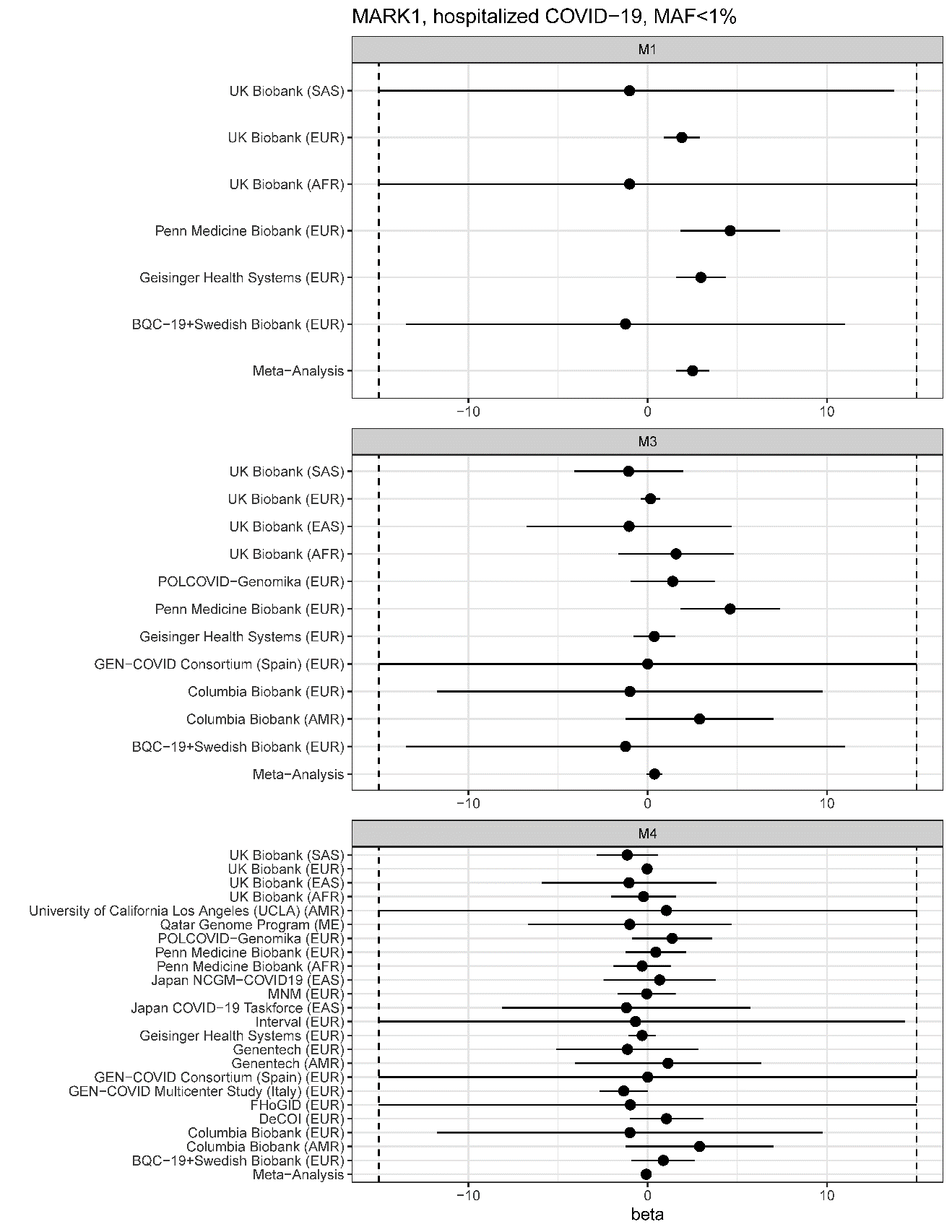

**Supplemental Figure 12**: *MARK1* beta coefficients (on logistic scale) with 95% interval for the hospitalized COVID-19 phenotype, MAF<1%. X-axes are cut at -10 and 10.

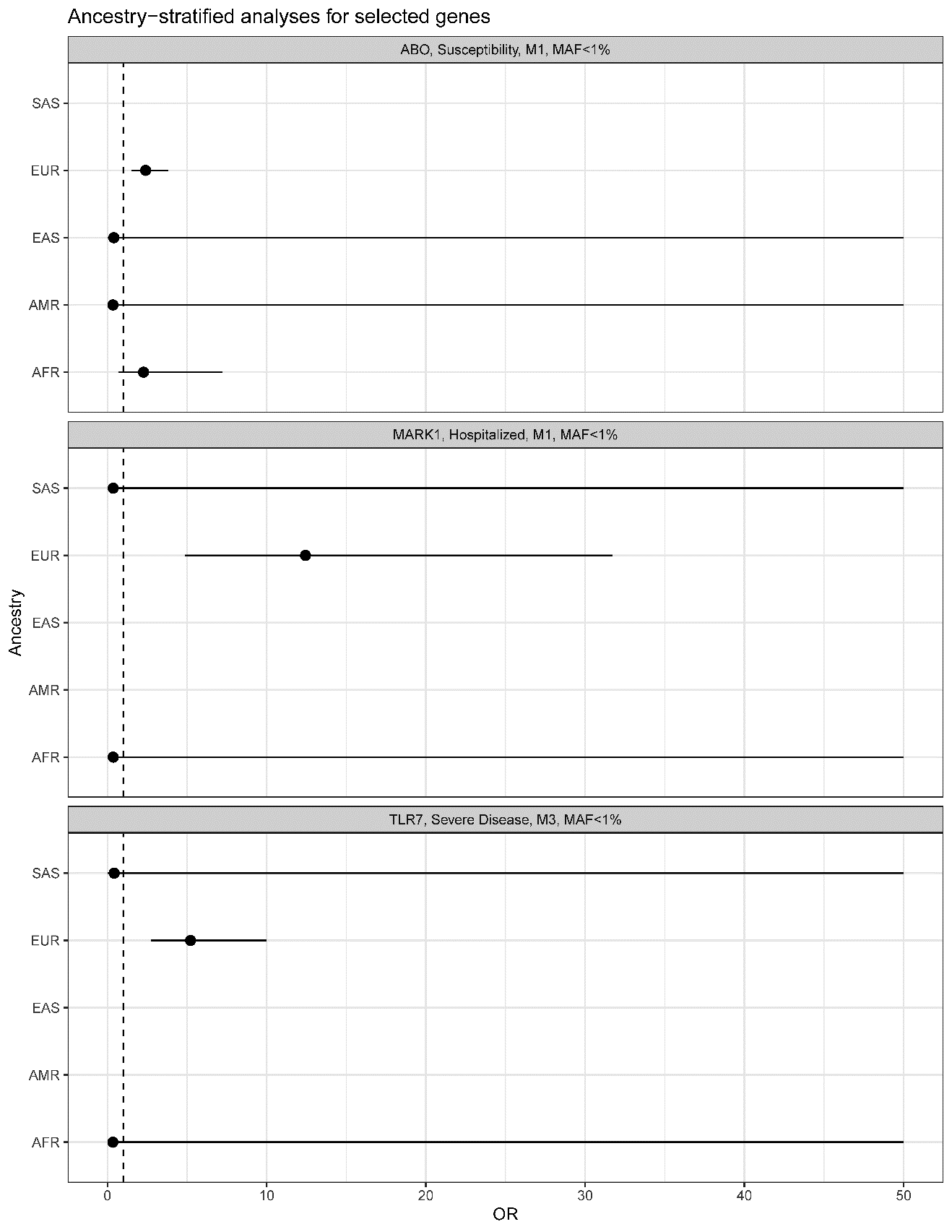

**Supplemental Figure 13**: Ancestry stratified results for *TLR7*, *MARK1*, and *ABO*. X-axis cut at 50. Figures show odds ratios and 95% confidence intervals
